# Magnetic susceptibility is predictive of future clinical severity in Parkinson’s disease

**DOI:** 10.1101/2023.07.12.23292555

**Authors:** George EC Thomas, Naomi Hannaway, Angelika Zarkali, Karin Shmueli, Rimona S Weil

**Affiliations:** Dementia Research Centre, UCL Institute of Neurology, London, UK; Department of Medical Physics and Biomedical Engineering, UCL, London, UK; Wellcome Centre for Human Neuroimaging, UCL, London, UK; Movement Disorders Consortium, UCL, London, UK

**Keywords:** Parkinson’s, dementia, MRI, Susceptibility, QSM, longitudinal

## Abstract

Dementia is common in Parkinson’s disease, but there is wide variation in its timing. With the emergence of disease modifying treatments in neurodegeneration, there is increasing need to detect and treat disease at the earliest stages and to monitor disease progression. A critical gap in Parkinson’s disease is the lack of quantifiable markers of progression, and methods to identify early stages. Until recently, MRI has had limited sensitivity to detect or track changes relating to Parkinson’s dementia, but advanced techniques, especially quantitative magnetic susceptibility mapping (QSM) which is sensitive to brain tissue iron, show potential for these purposes. So far, QSM, applied to cognition in Parkinson’s, has only been investigated in detail in cross-sectional cohorts. Here we present a longitudinal study throughout the brain using QSM in Parkinson’s disease. We included 59 Parkinson’s patients (with no dementia at study onset), and 22 controls, at baseline and after 3-year follow-up. Participants underwent detailed assessments of motor and cognitive severity at both timepoints. We found that increased magnetic susceptibility values in right temporal cortex and right putamen in patients were associated with poorer cognition at baseline. Strikingly, increased baseline susceptibility values within right temporal cortex, nucleus basalis of Meynert and putamen in patients were associated with greater cognitive severity after 3-year follow-up; and increased baseline susceptibility in basal ganglia, substantia nigra, red nucleus, insular cortex and dentate nucleus in patients were associated with greater motor severity after 3-year follow-up. We further found that after 3-years, increased follow-up susceptibility in these regions was associated with increased follow-up cognitive and motor severity, with further involvement of hippocampus relating to cognitive severity. However, region of interest analyses revealed that, in cortical regions, increases in paramagnetic susceptibility relating to iron could not alone explain the associations with increased clinical severity that were observed; and, similar to other studies, we did not find consistent increases in susceptibility within the 3-year follow-up period. Our study suggests that QSM has predictive sensitivity to detect changes in clinical severity many months prior to overt cognitive involvement in Parkinson’s. However, it also suggests that susceptibility within brain tissue over time may relate to changes in tissue composition other than iron concentration, such as in the relative proportion of myelin. Whilst our findings open the door for QSM to detect patients at the earliest stages of Parkinson’s dementia, we will require additional tissue metrics to augment the interpretation QSM, such that it can be used robustly in clinical practice or to provide outcome measures in therapeutic trials for disease modifying treatments in neurodegeneration.

## Introduction

Although thought of as a movement disorder, Parkinson’s disease causes significant non-motor symptoms, of which dementia and cognitive changes are amongst the most common and distressing. Dementia is six times more common in Parkinson’s than in the general population^1^ and causes higher levels of frailty and economic burden than other causes of dementia^2^. With the emergence of potentially effective disease modifying therapies for neurodegeneration^3, 4^, there is increasing pressure to start treatment at earlier stages. Being able to detect changes relating to neurodegeneration in patients with Parkinson’s at the earliest stages of Parkinson’s dementia, will potentially enable these patients to be identified for initiation of treatment. Detecting and tracking changes with progression of disease could have important applications for measuring and monitoring outcomes in these patients. Until recently, neuroimaging using MRI has shown only limited potential in detecting the early stages of dementia in Parkinson’s disease or monitoring Parkinson’s disease progression, mainly because conventional approaches have been based on volume or thickness of cortical regions, which are relatively insensitive in such contexts^5^. Instead, measures sensitive to changes in tissue composition, are more likely to detect and track neurodegeneration in Parkinson’s disease. We have recently shown that quantitative susceptibility mapping (QSM) which shows increased tissue magnetic susceptibility with higher levels of brain tissue iron, displays increased susceptibilities in relevant brain regions in patients with poorer cognition and worse motor scores^6^, and other studies have similarly shown these relationships in cross-sectional datasets^7, 8^. Measuring brain tissue iron is of particular relevance in pathological progression in Parkinson’s disease, as it is well-established that iron accumulates in the basal ganglia and substantia nigra in Parkinson’s disease^9, 10^; and excess tissue iron causes an increase in reactive oxygen species^11^ that interact with pathological proteins including alpha-synuclein^12^ and beta-amyloid^13^. We recently established that brain vulnerability to neurodegeneration in Parkinson’s disease relates to regional differences in gene expression for genes relating to heavy metal metabolism, including iron^14^.

However, whether QSM can detect changes in brain regions in patients who will go on to develop cognitive change over time, has not yet been proven; and whether it can track changes relating to neurodegeneration longitudinally, at whole brain or region of interest level, is not yet known.

So far, there have been very few longitudinal QSM studies in Parkinson’s disease, with none specifically examining the early stages and progression of cognitive decline. Three studies have investigated changes in susceptibility in pre-selected regions of interest (ROIs) over time, with a primary focus on the substantia nigra (SN) and relationships with motor severity^15–17^, with inconsistent findings. Du et al. reported decreasing susceptibility in the SN pars reticulata, but not the pars compacta, over 18 months in late-stage Parkinson’s disease and found that the decrease correlated with greater motor severity^15^. Guan et al. also reported susceptibility decreases in Parkinson’s disease patients with motor asymmetry, in the SN, globus pallidus and red nucleus over a period of 18 months^17^. Conversely, Bergsland et al. reported increased susceptibility in the ventral SN over a course of 3 years in people with Parkinson’s disease relative to controls, and did not find any association with clinical measures^16^. Some inconsistencies in these studies may relate to methodological differences. The QSM reconstruction pipeline consists of a series of processing steps (comprising brain masking, phase unwrapping, background field removal and dipole inversion) that can vary depending on choices made at each stage, based on MRI acquisition parameters as well as patient group.

Here, we have applied an optimised longitudinal QSM pipeline to identify susceptibility changes relating to cognitive severity in people with Parkinson’s followed longitudinally for 3 years, using a voxel-wise whole brain approach as well as a region of interest analysis. We hypothesised that increased susceptibilities in relevant brain regions at baseline would be associated with poorer future cognitive and motor performance in people with Parkinson’s disease; and that observed relationships between susceptibility and clinical severity would be stronger at follow-up than at the initial baseline timepoint. We also hypothesised that there would be an overall increase in QSM values during the disease course in Parkinson’s disease (beyond what would be expected with normal aging), particularly in iron rich deep-brain nuclei including the substantia nigra and basal ganglia.

## Materials and Methods

### Participants

107 patients with Parkinson’s disease (PD) who were within 10 years of diagnosis were recruited to this study between October 2017 and December 2018. They have been described previously^6^. Inclusion criteria were clinically diagnosed, early to mid-stage PD (Queen Square Brain Bank Criteria) aged 49-80 years. Exclusion criteria were confounding neurological or psychiatric disorders, dementia and metallic implants considered unsafe for MRI. Participants continued their usual therapy (including Levodopa) for all assessments and scans. No patients were taking cholinesterase inhibitors. In addition, 37 age-matched controls were recruited from sources including unaffected patient spouses. Participants were invited back three years after their initial visit with a follow-up interval of 38.5 ± 4.4 months (mean ± SD). Of the 144 (107 Parkinson’s) participants seen at baseline, 89 (67 Parkinson’s) were available and eligible at follow-up. After quality control of both susceptibility maps and MPRAGE images (by a rater blind to clinical outcomes), 8 Parkinson’s participants were excluded. This resulted in 81 (59 Parkinson’s) participants being included in the analyses described in this paper. A flowchart detailing reasons for exclusion from baseline analysis to follow-up can be seen in Figure 1. All participants gave written informed consent, and the study was approved by the Queen Square Research Ethics Committee. Patients with Parkinson’s were aged 63.0 ± 7.2 (mean ± SD), 28 females; controls were aged 67 ± 8.7, 12 females.

**Figure 1.**
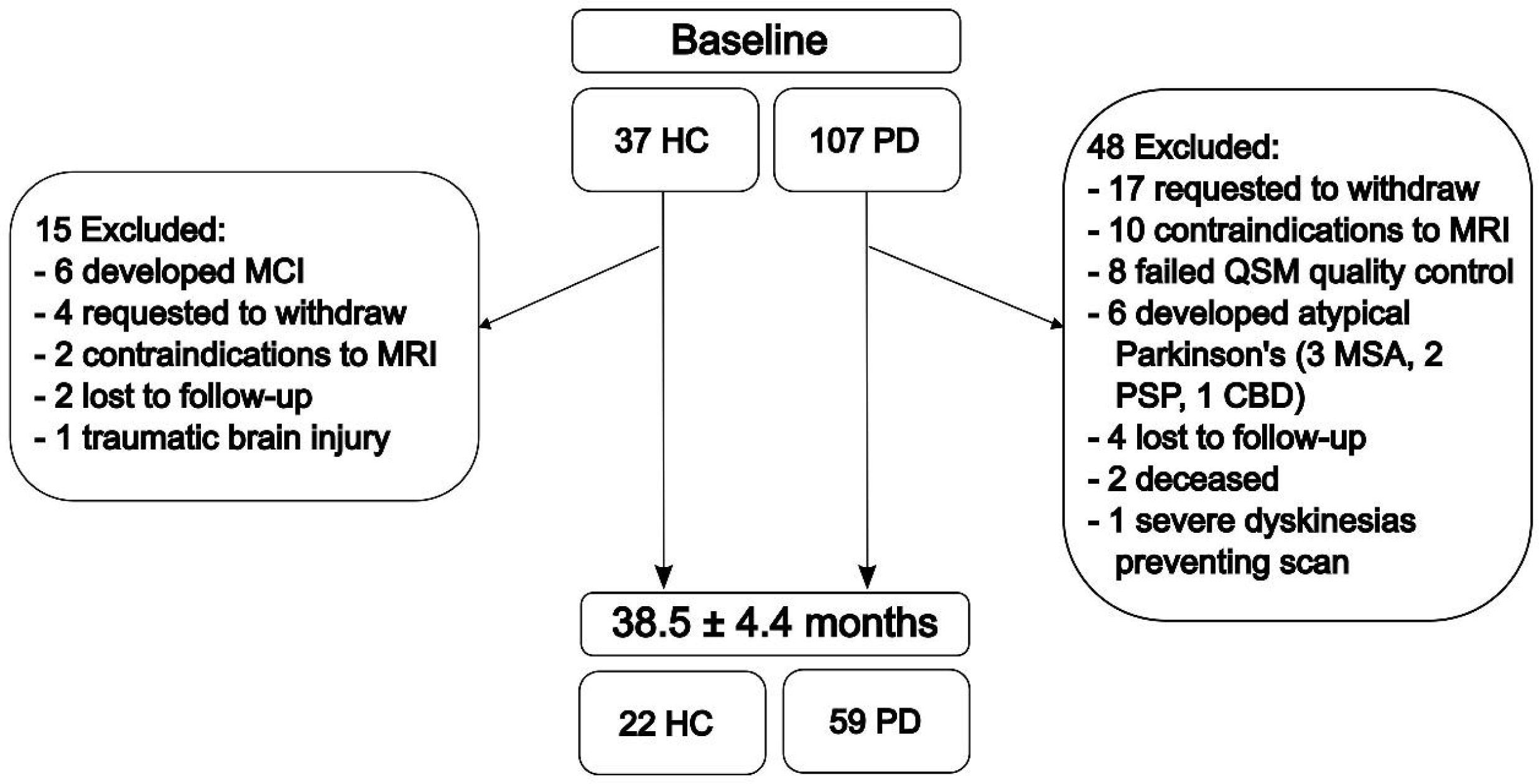
– Flowchart indicating participants involved in the study. Parkinson’s disease patients and controls tested in the study at baseline and after 36-month follow-up. Reasons for exclusion and data loss between baseline and follow-up visits are indicated. CBD = corticobasal degeneration; MSA = multiple system atrophy; PSP = progressive supranuclear palsy; QSM = quantitative susceptibility mapping.

### Clinical Assessments

All participants underwent clinical assessments at both timepoints comprising tests of motor function and cognition, and collection of clinical information. Detailed neuropsychological evaluation was performed as described previously^18^, with assessments in five cognitive domains. These were combined into a composite cognitive score using z-scores (based on control means and standard deviations) for tests across these five cognitive domains (Stroop colour, Hooper visual organisation, word recognition, verbal fluency category, verbal fluency letter), as we have previously described^19^. As this composite cognitive score is based on a more detailed cognitive assessment, it is likely to have greater variance and greater sensitivity to differences between individuals than short global cognitive assessments such as the Montreal Cognitive Assessment (MoCA). Motor function was assessed using the Movement Disorder Society Unified Parkinson’s Disease Rating Scale part III (MDS-UPDRS III) with patients in the ‘On’ state. Additional tests including smell, depression and anxiety, and REM sleep behavioural disorder questionnaires were also carried out. Participant demographics and clinical measures for both groups at baseline and follow-up are shown in Table 1.

**Table 1.** Demographics table for participants included in longitudinal analysis. Means (SDs) reported. UPDRS-III = Unified Parkinson’s Disease Rating Scale part 3 (motor score). LEDD = Levodopa equivalent dose. HADS = Hospital Anxiety and Depression Scale. RBDSQ = REM (Rapid Eye Movement) Sleep Behaviour Disorder Screening Questionnaire. Statistical tests: OR = Fisher’s exact test odds ratio, T = two-tailed t-test test statistic, U = Mann-Whitney U-test test statistic.

| Measure | Controls<br>(N = 22) | Parkinson's<br>disease<br>(N = 59) | Stat | P |
| --- | --- | --- | --- | --- |
| Sex (F / M) | 12 / 10 | 28 / 31 | T = 1.33 | 0.62 |
| Age (years) |  |  |  |  |
| *Baseline* | 66.7 (8.7) | 63.0 (7.2) | T = 1.93 | 0.057 |
| *Follow-up* | 69.9 (8.8) | 66.2 (7.1) | T = 1.95 | 0.055 |
| Years of education | 18.0 (1.8) | 17.2 (2.7) | U = 994 | 0.32 |
| MoCA score |  |  |  |  |
| *Baseline* | 29.1 (1.1) | 28.4 (1.7) | U = 1054 | 0.094 |
| *Follow-up* | 29.0 (1.5) | 27.8 (2.8) | U = 1061 | 0.082 |
| Combined cognitive score |  |  |  |  |
| *Baseline* | 0.11 (0.57) | -0.11 (0.74) | U = 1004 | 0.28 |
| *Follow-up* | 0.07 (0.49) | -0.23 (0.91) | U = 1003 | 0.29 |
| UPDRS-III |  |  |  |  |
| *Baseline* | 6.1 (5.0) | 22.3 (12.3) | T = -6.01 | 5.4x10<sup>-8</sup> |
| *Follow-up* | 5.2 (4.2) | 28.7 (9.1) | U = 263.5 | 1.2x10<sup>-11</sup> |
| HADS depression |  |  |  |  |
| *Baseline* | 1.0 (1.2) | 3.4 (2.7) | U = 524.5 | 4.7x10<sup>-5</sup> |
| *Follow-up* | 2.0 (2.6) | 4.3 (2.7) | U = 565.5 | 3.2x10<sup>-4</sup> |
| HADS anxiety |  |  |  |  |
| *Baseline* | 3.5 (3.7) | 5.8 (3.9) | U = 666.5 | 0.012 |
| *Follow-up* | 3.6 (3.8) | 5.8 (3.9) | U = 640 | 0.0052 |
| RBDSQ score |  |  |  |  |
| *Baseline* | 2.3 (1.4) | 3.8 (2.3) | U = 669 | 0.012 |
| *Follow-up* | 1.6 (1.5) | 4.7 (3.0) | U = 468.5 | 3.6x10<sup>-6</sup> |
| Disease duration (years) |  |  |  |  |
| *Baseline* | - | 4.1 (2.6) | - | - |
| *Follow-up* | - | 7.3 (2.7) | - | - |
| LEDD (mg) |  |  |  |  |
| *Baseline* | - | 432.1 (228.0) | - | - |
| *Follow-up* | - | 651.2 (354.7) | - | - |
| Motor deficit dominance<br>(left / right / both) |  |  |  |  |
| *Baseline* | - | 23 / 32 / 4 | - | - |
| *Follow-up* | - | 24 / 27 / 8 | - | - |

### MRI protocol

At both timepoints, MRI measurements consisting of susceptibility- and T1-weighted scans were performed on the same Siemens Prisma-fit 3T MRI system using a 64-channel receive array coil (Siemens Healthcare, Erlangen, Germany). Susceptibility-weighted MRI signals were obtained from a 2×1-accelerated^20^, 3D flow-compensated spoiled-gradient-recalled echo sequence. Flip angle=12°; echo time=18 ms; repetition time=25 ms; receiver bandwidth=110 Hz/pixel; matrix size=204×224×160 with 1×1×1 mm^3^ voxel size; scan time= 5 min 41 s. T1-weighted magnetization-prepared 3D rapid gradient-echo (MPRAGE) anatomical images were acquired with the following parameters: 2×1 parallel acceleration; inversion time=1100 ms; flip angle=7°; first echo time=3.34 ms; echo spacing=7.4 ms; repetition time=2530 ms; receiver bandwidth=200 Hz/pixel; matrix dimensions=256×256×176; voxel size=1×1×1 mm^3^; scan time=6 min 3 s. Follow-up scans were performed after an average of 38.5 ± 4.4 months (mean ± SD).

### QSM reconstruction

3D complex phase data (multi-channel coil data were adaptively combined using scanner software) were unwrapped using a rapid path-based based minimum spanning tree algorithm (ROMEO)^21^. Brain masks were calculated from magnitude images using FSL’s brain extraction tool (BET2)^22^ iteratively for robust centre-of-gravity estimation. Prior to background field removal a noise map was estimated from inverted magnitude images and thresholded to allow noise-based exclusion to be carried out on the outer 3 layers of voxels^23^. This was done to remove voxels with a low signal-to-noise ratio likely to contain unreliable phase data. Background field removal was completed in a two-step process; first using Laplacian boundary value extraction^24^, followed by 3D polynomial residual fitting and removal^25^, revealing the local field map. Prior to dipole inversion, one voxel was eroded from the edge of the brain mask. Dipole inversion was completed using multi-scale dipole inversion (MSDI)^26^ to estimate the underlying susceptibility. All image processing was carried out using MATLAB (The MathWorks, Inc., Natick, MA, USA) unless specified otherwise. After susceptibility maps had been calculated, both the baseline and follow-up maps for all 89 subjects were visually inspected for quality control purposes, by a rater blind to clinical outcomes. After this, seven subjects were excluded due to erroneous susceptibility maps (either at baseline or at follow-up) due to excessively noisy phase data or the presence of artefacts. This resulted in 81 (59 Parkinson’s) participants being included in subsequent analyses.

### Spatial standardization

For spatial standardization of susceptibility maps, we used the same method as described previously^6^. The previously optimised^27^ methodology involved creation of an average T1-weighted template space from MPRAGE images, processed using the default segmentation pipeline in SPM12 (https://www.fil.ion.ucl.ac.uk/spm/software/spm12/), from all participants. This was modified for follow-up analysis, in that both baseline and follow-up T1 images were used to produce the template, rather than just baseline images. As before, N4ITK bias-field-corrected^28^ magnitude images were rigidly, then affinely co-registered to their corresponding MPRAGE volumes, and QSM spatial standardization was achieved through applying the above transformations, concatenated to co-register each QSM to its corresponding MPRAGE and then to the average T1-weighted template, using high-order b-spline interpolation. All QSM images co-registered to the template were visually inspected to ensure that there were no errors in the applied transformation.

### Whole-brain QSM statistical analysis

Voxel-wise statistical analyses of QSM throughout the brain used a similar protocol to that described previously^6^ and was performed using absolute QSM data. This is required for statistical conditioning in whole brain QSM analyses due to the close proximity of positive and negative QSM values that may otherwise cancel each other out during spatial smoothing when signed QSM data are used^29^. To attenuate the impact of misregistration and other inaccuracies, images were spatially smoothed using a 3D Gaussian kernel (3-mm standard deviation), and were subsequently smoothing compensated^29^. Permutation analyses were performed with Randomise v2.9 and threshold-free cluster enhancement (http://fsl.fmrib.ox.ac.uk/fsl/fslwiki/Randomise) in FSL. Significant clusters across the whole brain were inferred from a random subset of 10,000 data permutations and reported at family-wise error-corrected p<0.05 (P_FWE_ < 0.05). Design and contrast matrices for the permutation analyses were generated using FSL’s GLM function (https://fsl.fmrib.ox.ac.uk/fsl/fslwiki/GLM).

The following permutation analyses were then performed. Firstly, a set of cross-sectional analyses were run to investigate differences in magnetic susceptibility between the control and Parkinson’s groups at both baseline and at follow-up, adjusting for age and sex. To investigate within-group change in susceptibility between visits in controls and Parkinson’s, single-group paired t-tests adjusted for age, sex, and time between scans were carried out.

To investigate how susceptibility relates to motor and cognitive severity over time, regression analyses were performed for 1) baseline susceptibility against baseline MDS-UPDRS-III, and combined cognitive score (adjusted for age at baseline and sex); 2) baseline susceptibility against follow-up MDS-UPDRS-III, and combined cognitive score (adjusted for age at baseline, sex and time between scans); and 3) follow-up susceptibility against follow-up MDS-UPDRS-III, and combined cognitive score (adjusted for age at follow-up and sex). Given the primary interest in relating QSM to the clinical severity of Parkinson’s, all regression analyses were performed using only people with Parkinson’s disease. However, they were additionally run in the control group as a point of reference.

After analysis, the QSM template and statistical maps were transformed into MNI152 space (Montreal Neurological Institute, McGill University, Canada) for display purposes, using a previously optimised co-registration approach^27^.

### Regional QSM statistical analysis

*Post-hoc* region-of-interest (ROI) analyses were additionally carried out to probe the nature of significant interactions observed between motor and cognitive severity and absolute, as well as signed susceptibility in the PD group. Use of signed susceptibility allowed us to specifically examine each brain region to determine whether relationships observed as significant in whole brain analyses were likely due to diamagnetic or paramagnetic sources of susceptibility. Use of absolute susceptibility served as a point of comparison, and as confirmation for the whole brain results. Therefore, the following ROIs were selected based on the results of the whole brain analysis: substantia nigra pars compacta (SNpc) and pars reticulata (SNpr), red nucleus, caudate nucleus, putamen, globus pallidus, hippocampus, nucleus basalis of Meynert (NBM), insular cortex, lateral and medial orbitofrontal cortex, medial orbitofrontal cortex, and dentate nucleus. The caudate nucleus, globus pallidus, putamen and hippocampus were segmented automatically using FSL-FIRST on the study-wise MPRAGE template. As the red nucleus is not included in FSL-FIRST, it was segmented from the MPRAGE and QSM templates using the MRIcloud^30^ QSM segmentation tool^31^. MRIcloud does not subdivide the SNpc and SNpr so these were manually traced on the MPRAGE template, where T1 shortening effects exhibited by the neuromelanin-rich neurons of the SNpc are visible in comparison to the SNpr^27^. As it was not segmented either by FSL-FIRST or MRIcloud, the NBM was manually traced as described previously^6^. All subcortical regions of interest were eroded by convolution with a 1-mm radius spherical kernel to minimise partial-volume effects from co-registration discrepancies. The insula, lateral and medial orbitofrontal cortices were defined using the Desikan-Killiany-Tourville digital atlas. The OASIS-30 template and OASIS-TRT20 joint fusion atlas were obtained from Mindboggle (mindboggle.info/data.html). The study-wise-template to OASIS-30 space non-linear transformations were calculated as described previously^27^. To reduce partial-volume contamination, each cortical region of interest was intersected with a study-wise average grey matter mask (generated in SPM12) binarized at a grey matter density cut-off of 0.5 using FSL. See Supplementary Figure 1 the ROIs visually presented on the QSM template in MNI space. Unsmoothed, mean absolute and signed susceptibility values were extracted from all above ROIs and values were averaged across hemispheres to improve measurement stability. All analyses were performed in R version 4.4.2, using the ‘lm’ function for linear models, and the ‘lme4’ package for linear mixed models (https://CRAN.R-project.org/package=lme4).

The strength of relationships between susceptibility and clinical severity at baseline and follow-up were examined by fitting linear models at each ROI as follows: *QSM*(*ROI*)_*ik*_ = *β*_0_ + *β*_1_*Age*_*ik*_ + *β*_2_*Sex*_*i*_ + *β*_3_*clin*_*ik*_ + ε_*ik*_, where *QSM*(*ROI*)_*ik*_ is the regional mean absolute or signed susceptibility at timepoint *k* for subject *i*, *clin*_*ik*_ is either the combined cognitive or MDS-UPDRS-III score for participant *i* at timepoint *k*, *Age*_*ik*_ is the age of participant *i* at timepoint *k*, and *Sex*_*i*_ is the sex of participant *i*. In all linear and linear mixed models, *β* and ε are the model coefficients and fit residuals, respectively. To account for multiple comparisons, p-values were FDR adjusted across the 12 ROIs^32^. The relationship between susceptibility and future motor and cognitive severity was tested as follows: *QSM*(*ROI*)_*ik*1_ = *β*_0_ + *β*_1_*Age*_*ik*1_ + *β*_2_*Sex*_*i*_ + *β*_3_*Tfup*_*i*_ + *β*_4_*clin*_*ik*2_ + ε_*i*_, where *QSM*(*ROI*)_*ik*1_ is the regional mean absolute or signed susceptibility at baseline for participant *i*, *clin*_*ik*2_is either the combined cognitive or MDS-UPDRS-III score for participant *i* at follow-up, and *Age*_*ik*1_, *Tfup*_*i*_ and *Sex*_*i*_ are the baseline age, time between scans and sex for participant *i*, respectively. ANOVA tests were used to determine significance and relevant test-statistics for each model.

To facilitate comparisons to previous longitudinal QSM studies in Parkinson’s disease, a set of ROI analyses to look at the change in the *signed* susceptibilities in Parkinson’s disease over time were carried out. To do this, the following linear mixed model was fitted at each ROI: *QSM*(*ROI*)_*ik*_ = *β*_0_ + *β*_1_*Age*_*ik*1_ + *β*_2_*Sex*_*i*_ + *β*_3_*Tfup*_*ik*_ + *b*_0*i*_ + ε_*ik*_, where *QSM*(*ROI*)_*ik*_is the regional mean signed susceptibility at timepoint *k* for participant *i*, *Tfup*_*ik*_is the time from baseline at timepoint *k* for participant *i* (this will be zero at baseline), *Age*_*ik*1_ and *Sex*_*i*_are the age at baseline and sex for participant *i*, and *b*_0*i*_ is a random intercept effect per subject.

Finally, in line with previous studies^33^, magnetic susceptibility in the SNpc and SNpr was compared between controls and people with Parkinson’s disease at both baseline and at follow-up. The following linear models were fitted in the SNpc and SNpr: *QSM*(*ROI*)_*ik*_ = *β*_0_ + *β*_1_*Age*_*ik*1_ + *β*_2_*Sex*_*i*_ + *β*_3_*PD*_*i*_ + ε_*ik*_, where *QSM*(*ROI*)_*ik*_ is the regional mean absolute or signed susceptibility at timepoint *k* for participant *i*, *PD*_*i*_ is the control or Parkinson’s disease status of participant *i*, *Age*_*ik*_ is the age of participant *i* at timepoint *k*, and *Sex*_*i*_ is the sex of participant *i*.

### Voxel based morphometry

To compare the performance of QSM and conventional atrophy-based measures in predicting clinical severity, whole brain voxel-based morphometry (VBM) analyses mirroring the whole brain QSM analyses were also carried out. Segmentation, normalisation to Montreal Neurological Institute (MNI) space and tissue probability modulation were carried out in SPM12 (http://www.fil.ion.ucl.ac.uk/spm/software/spm12) with default parameters, in conjunction with the DARTEL toolbox using a Gaussian smoothing kernel of 8 mm full-width-at-half-maximum. Images were compared cross-sectionally between patients and controls using two-sample t-tests, and multiple regression models were implemented to examine associations between voxel-wise grey and white matter volume and clinical parameters (combined cognitive score and UPDRS-III). All models included age, sex and total intracranial volume as nuisance covariates, and regression models examining baseline volume in relation to follow-up clinical scores were additionally adjusted for time between scans. Statistical parametric maps were generated for all models and reported at P^FWE^ < 0.05.

## Results

### Association between magnetic susceptibility and clinical severity over time

We examined three associations: 1) baseline QSM with baseline clinical measures (for associations already present at baseline); 2) baseline QSM with follow-up clinical measures (for QSM *predicting* clinical outcomes); and 3) follow-up QSM with follow-up clinical measures (to examine strength of associations after follow-up). It should be noted that *lower* combined cognitive scores reflect poorer cognitive ability, whereas *higher* UPDRS-III scores reflect poorer motor ability.

In people with Parkinson’s disease, there was a significant association between increased baseline absolute susceptibility and decreased baseline cognitive score in the right temporal cortex, with a small number of significant voxels in the right putamen (Figure 2A, P_FWE_ < 0.05). 2) After clinical follow-up, we found a significant relationship between increased baseline absolute susceptibility and decreased follow-up cognitive score in the right temporal cortex, right basal forebrain, and right putamen (Figure 2B, P_FWE_ < 0.05). 3) Finally, we examined increased follow-up susceptibility and its relationship with decreased follow-up cognitive performance, where we observed more widespread increases in QSM, with significant clusters bilaterally in temporal regions including hippocampi, brainstem regions including the red nucleus as well as right putamen, basal forebrain and insular cortex (Figure 2C, P_FWE_ < 0.05). In all three analyses the opposite contrasts (absolute QSM increasing with increasing cognitive score) revealed no significant clusters at P_FWE_ < 0.05.

**Figure 2.**
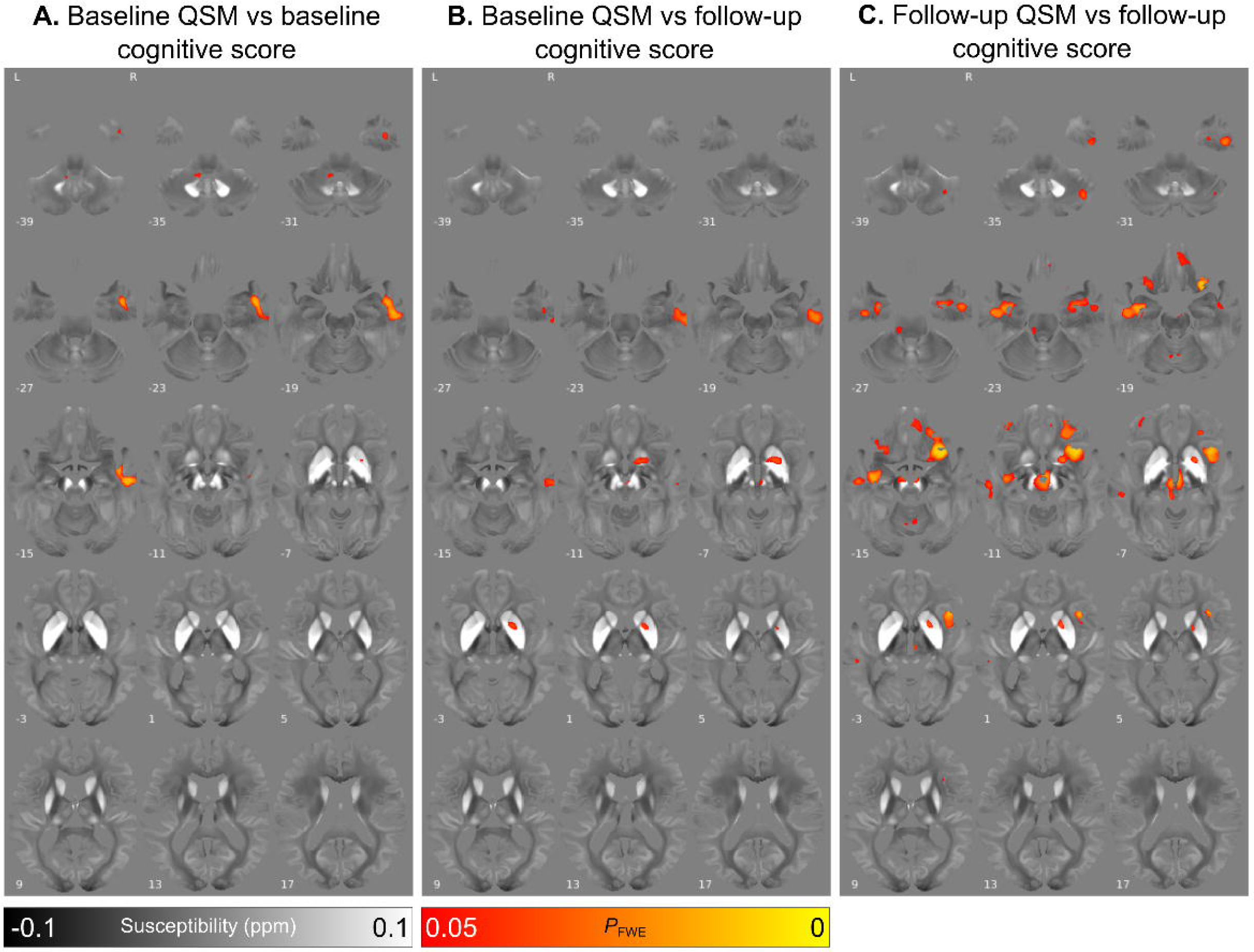
- Relationship between increased absolute magnetic susceptibility and declining cognitive ability over the Parkinson’s disease course, in a whole brain analysis. **A.** Association between baseline susceptibility and composite cognitive score at baseline, adjusted for age at baseline and sex. **B**. Association between baseline susceptibility and composite cognitive score at 3-year follow-up, adjusted for age at baseline, sex and time between scans. **C.** Association between susceptibility at 36-month follow-up and composite cognitive score at 3-year follow-up, adjusted for age at follow-up and sex. Results are overlaid on the study-wise QSM template in MNI space, and numbers represent axial slice location in MNI space. Left side is shown on the left. Red/yellow clusters represent voxels where a significant relationship was seen at FWE-corrected P<0.05.

### ROI analyses to further explore the relationship between QSM values and clinical severity

Post-hoc ROI analyses of signed susceptibility mirroring the three whole brain analyses were carried out to specifically examine the relative contribution of para- and diamagnetic susceptibility sources. At baseline (see Supplementary Figure 2), there was a significant negative association between cognition and absolute susceptibility in the lateral orbitofrontal cortex (P_FDR_ = 0.029, β = -2.2x10^-3^). Other associations were observed in the red nucleus and caudate nucleus for both absolute and signed susceptibility, but these were not significant after correction for multiple comparisons.

For the relationship between baseline susceptibility and follow-up cognition (see Figure 3), there were significant negative associations between follow-up cognition and both absolute and signed susceptibility in the SNpr (absolute/signed P_FDR_ = 0.032/0.032, β = -9.6x10^-3^/- 1.0x10^-2^), red nucleus (P_FDR_ = 0.026/0.028, β = -7.8x10^-3^/-7.8x10^-3^), NBM (P_FDR_ = 0.024/0.028, β = -1.8x10^-2^/-1.8x10^-2^), caudate nucleus (P_FDR_ = 0.024/0.028, β = -5.4x10^-3^/- 5.3x10^-3^) and putamen (P_FDR_ = 0.026/0.028, β = -7.3x10^-2^/-7.4x10^-2^). For subcortical regions, mean absolute and signed susceptibilities were almost identical (and overlap in Figure 3). However, in cortical structures, notably, hippocampi, insular cortex and lateral and medial orbitofrontal cortex, mean absolute and signed susceptibilities diverged, suggesting additional effects of diamagnetic sources in these regions on cognitive scores (and see non-overlapping values in Figure 3).

**Figure 3.**
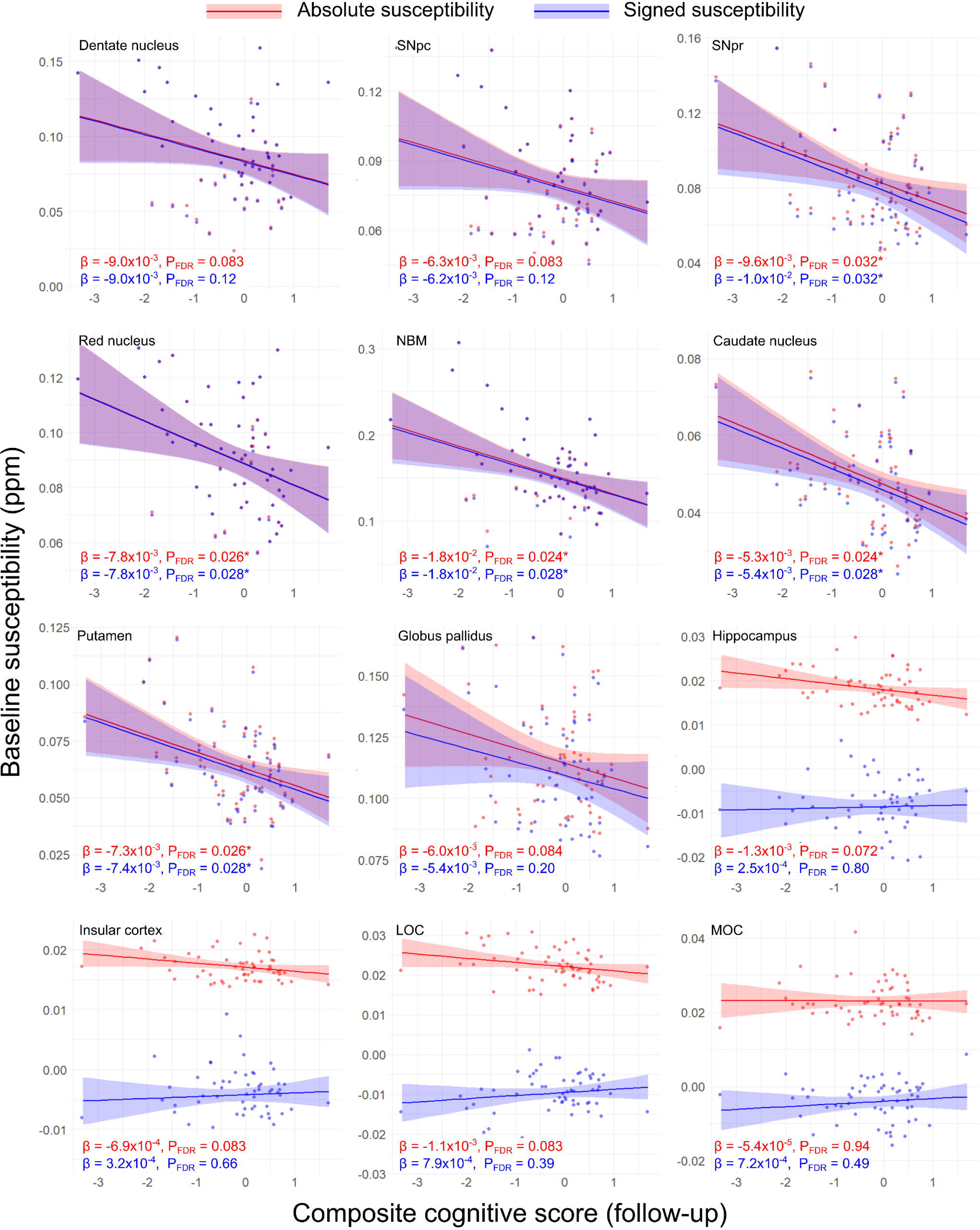
– Regional relationships between baseline magnetic susceptibility and follow-up cognitive score in Parkinson’s disease. Data and statistics relating to ROI mean absolute susceptibility are shown in red, and those relating to ROI mean signed susceptibility are shown in blue. Results are adjusted for age at baseline, sex and time between scans. FDR-corrected p-values (P_FDR_) are presented, with asterisks indicating significant interactions at P_FDR_ < 0.05. β is the linear model coefficient associated with composite cognitive score. SNpc/pr = substantia nigra pars compacta / pars reticulata; NBM = nucleus basalis of Meynert; MOC = medial orbitofrontal cortex; LOC = lateral orbitofrontal cortex.

At follow-up (see Supplementary Figure 3), there were significant negative associations between cognition and absolute and signed susceptibilities in the SNpc (absolute/signed P_FDR_ = 0.015/0.030, β = -9.2x10^-3^/-9.4x10^-3^), red nucleus (P_FDR_ = 0.0079/0.016, β = -1.0x10^-2^/- 1.0x10^-2^), NBM (P_FDR_ = 0.015/0.030, β = -1.6x10^-2^/-1.6x10^-2^), and caudate nucleus (P_FDR_ = 0.015/0.030, β = -5.4x10^-3^/-5.5x10^-3^). In the SNpr there was a significant negative association between cognition and absolute susceptibility (P_FDR_ = 0.047, β = -8.3x10^-3^), although the effect on signed susceptibility was not significant after correction for multiple comparisons. Interestingly, in the cortical ROIs the relationship between cognition and absolute and signed susceptibility diverged. There were significant positive associations observed between cognition and absolute susceptibility in the insular cortex (absolute/signed P_FDR_ = 0.0079/0.046, β = -1.2x10^-3^/1.4x10^-3^), and lateral orbitofrontal cortex (P_FDR_ = 0.015/0.030, β = -1.7x10^-3^/1.7x10^-3^). There were also significant negative associations between cognition and absolute susceptibility in the hippocampus (P_FDR_ = 0.015, β = -1.9x10^-3^), and medial occipital cortex (P_FDR_ = 0.015, β = -2.0x10^-3^). However, for signed QSM, there was a positive association with cognition (rather than negative as had been found for absolute values) seen in the hippocampi, insular and lateral orbitofrontal cortices, although these were not significant after correction for multiple comparisons in the hippocampi. Full ROI statistics for the relationship between susceptibility and cognition can be seen in Supplementary Table 1.

With regards to motor severity, no significant clusters were found for increased baseline absolute susceptibility correlating with increased baseline MDS-UPDRS III score (Figure 4A). However, there were widespread regions found showing associations between increased baseline absolute susceptibility and increased follow-up MDS-UPDRS III score including: bilaterally in the basal-ganglia, substantia nigra, red nucleus and insular cortex, as well as right dentate nucleus (Figure 4B, P_FWE_ < 0.05). The relationship between increased follow-up absolute susceptibility and increased follow-up MDS-UPDRS III score followed a similar pattern of regional involvement (Figure 4C, P_FWE_ < 0.05). In all three analyses the opposite contrasts (absolute susceptibility increasing with decreasing MDS-UPDRS III score) revealed no significant clusters at P_FWE_ < 0.05.

**Figure 4.**
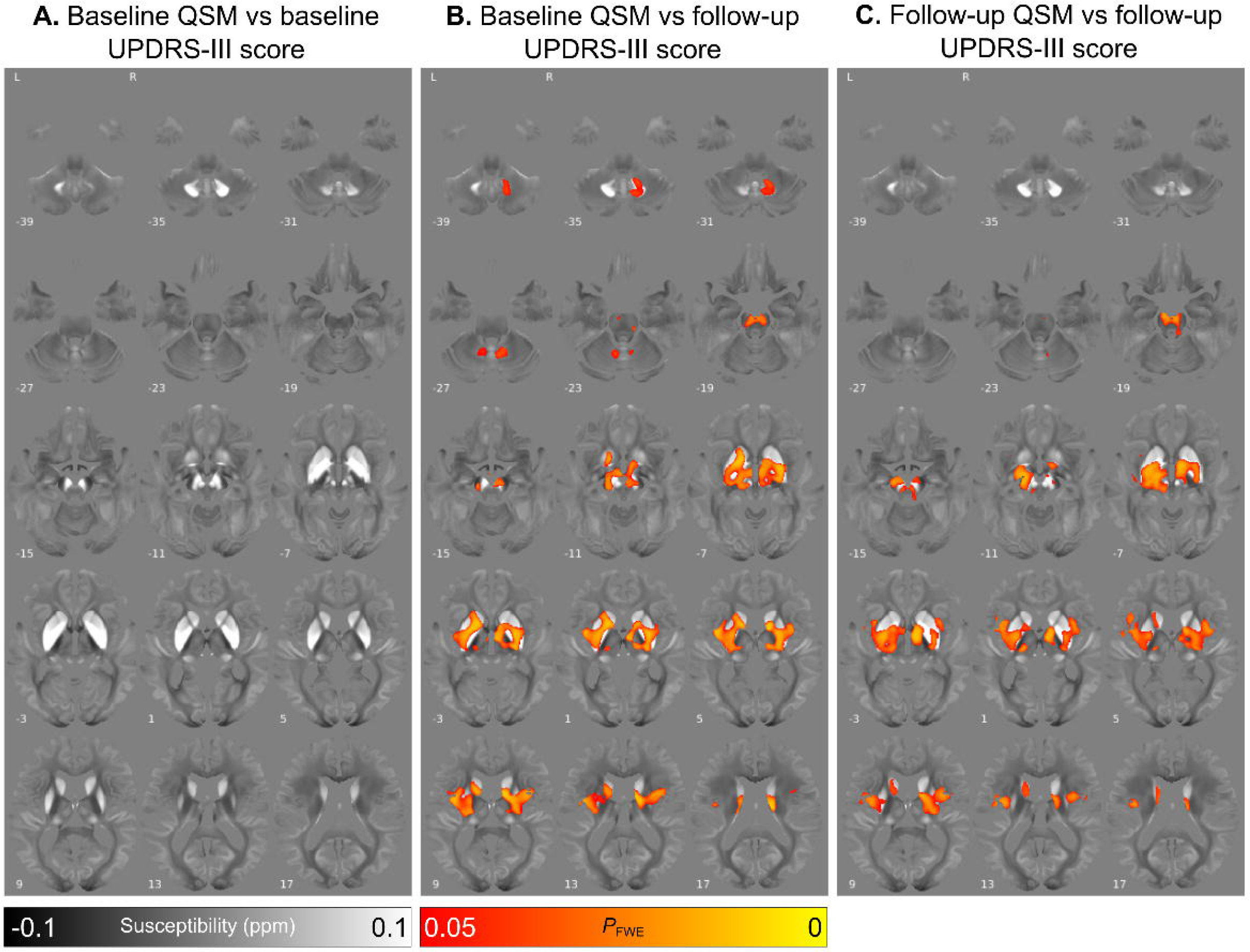
- Relationship between increased absolute magnetic susceptibility and increasing motor severity over the Parkinson’s disease course, in a whole brain analysis. **A.** Association between baseline susceptibility and MDS-UPDRS III score at baseline, adjusted for age at baseline and sex. **B.** Association between baseline susceptibility and MDS-UPDRS III score at 3-year follow-up, adjusted for age at baseline, sex and time between scans. **C.** Association between susceptibility at 3-year follow-up and MDS-UPDRS III score at 3-year follow-up, adjusted for age at follow-up and sex. Results are overlaid on the study-wise QSM template in MNI space, and numbers represent axial slice location in MNI space. Left side is shown on the left. Red/yellow clusters represent voxels where a significant relationship was seen at FWE-corrected P<0.05.

At ROI level, there were no significant relationships between baseline MDS-UPDRS-III score and baseline absolute or signed susceptibility (see Supplementary Figure 4). Looking at baseline susceptibility versus follow-up MDS-UPDRS III score (see Figure 5), there were significant positive associations with both absolute and signed susceptibility in the dentate nucleus (absolute/signed P_FDR_ = 0.011/0.010, β = 1.6x10^-3^/1.6x10^-3^), SNpr (P_FDR_ = 0.031/0.034, β = 9.4x10^-4^/9.8x10^-4^), red nucleus (P_FDR_ = 0.012/0.011, β = 9.3x10^-4^/9.4x10^-4^), NBM (P_FDR_ = 0.012/0.014, β = 1.9x10^-3^/2.0x10^-3^), caudate nucleus (P_FDR_ = 0.031/0.033, β = 4.6x10^-4^/4.7x10^-4^), putamen (P_FDR_ = 0.027/0.027, β = 7.2x10^-4^/7.2x10^-4^) and globus pallidus (P_FDR_ = 0.031/0.046, β = 8.2x10^-4^/7.9x10^-4^). Similar to the ROI analysis for QSM data and cognition, absolute and signed QSM data were almost overlapping for subcortical regions but showed differences in cortical regions. A positive association was observed with absolute susceptibility in the insula although this was not significant after correction for multiple comparisons, whereas a significant negative association was found between signed susceptibility and MDS-UPDRS III score in the insular cortex (P_FDR_ = 0.027, β = -1.6x10^-4^).

**Figure 5.**
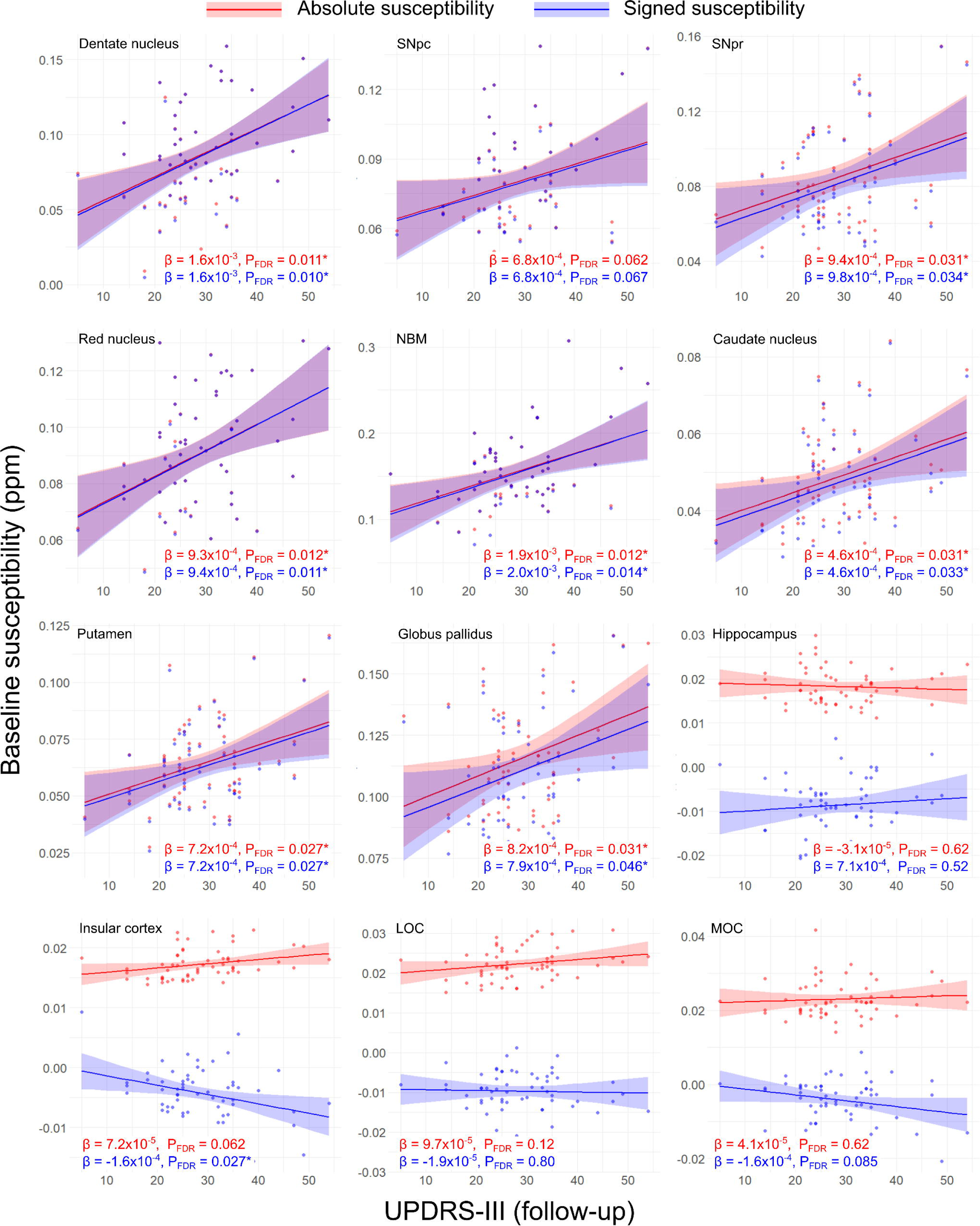
– Regional relationships between baseline magnetic susceptibility and follow-up motor severity score in PD. Data and statistics relating to absolute magnetic susceptibility are shown in red, and those relating to signed susceptibility are shown in blue. Results are adjusted for age at baseline, sex and time between scans. FDR-corrected p-values (P_FDR_) are presented, with asterisks indicating significant interactions at P_FDR_ < 0.05. β is the linear model coefficient associated with MDS-UPDRS-III score. SNpc/pr = substantia nigra pars compacta / pars reticulata; NBM = nucleus basalis of Meynert; MOC = medial orbitofrontal cortex; LOC = lateral orbitofrontal cortex; MDS-UPDRS-III = Movement Disorders Society Unified Parkinson’s Disease Rating Scale part III.

At follow-up (see Supplementary Figure 5), there were significant positive associations between MDS-UPDRS III score and both signed and absolute susceptibilities in the dentate nucleus (absolute/signed P_FDR_ = 0.013/0.014, β = 1.6x10^-3^/1.6x10^-3^), SNpr (P_FDR_ = 0.013/0.014, β = 1.2x10^-3^/1.3x10^-3^), NBM (P_FDR_ = 0.013/0.014, β = 1.8x10^-3^/1.8x10^-3^), caudate nucleus (P_FDR_ = 0.036/0.041, β = 4.7x10^-4^/4.8x10^-8^) and putamen (P_FDR_ = 0.031/0.040, β = 7.5x10^-4^/7.4x10^-4^). There was also a significant positive association of MDS-UPDRS-III score with absolute susceptibility in the globus pallidus (P_FDR_ = 0.036, β = 7.9x10^-4^), although the association with signed susceptibility was not significant after correction for multiple comparisons. Similar to relationships seen with cognition, we found opposite directions of association in cortical ROIs, with a negative association in the insular cortex between absolute susceptibility and MDS-UPDRS III score (P_FDR_ = 0.036, β = 9.2x10^-5^), compared with a negative association with signed susceptibility (P_FDR_ = 0.041, β = - 1.5x10^-4^). Full ROI statistics for motor severity can be seen in Supplementary Table 2.

In the control group, at whole-brain level, there were no significant clusters observed for any associations between magnetic susceptibility and motor or cognitive scores.

### Changes in susceptibility over time

In people with Parkinson’s disease, absolute susceptibility was significantly increased at follow-up relative to baseline in the left precentral gyrus, left middle frontal cortex and right middle temporal gyrus (P_FWE_ < 0.05, Figure 6). Using an uncorrected threshold for significance (p<0.05), additional increases in absolute susceptibility were seen over time in the basal ganglia, frontal, parietal and temporal cortices and right substantia nigra, with decreases in the left dentate nucleus and posterior corpus callosum (see Supplementary Figure 6). In the control group, there were no significant clusters at whole brain analysis indicating differences between baseline and follow-up absolute susceptibility; and using an uncorrected threshold (p<0.05) some sparse cortical increases and decrease were observed, as well as some left mesencephalic and pallidal decreases (see Supplementary Figure 6).

**Figure 6.**
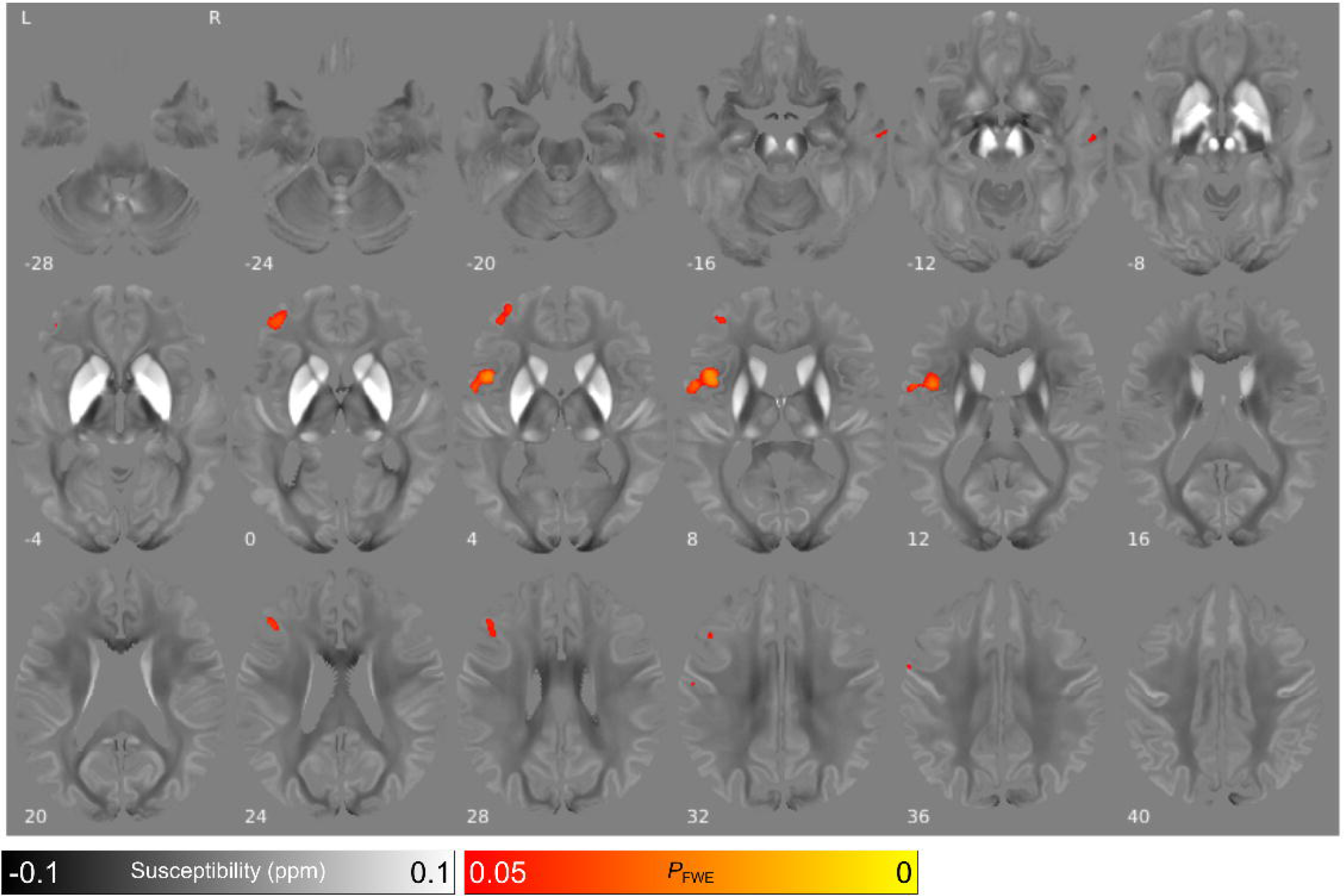
– Changes in absolute magnetic susceptibility over time in Parkinson’s disease at whole brain. Whole brain analysis is adjusted for age at baseline, sex and time between scans. Red/yellow clusters represent voxels where absolute QSM was significantly higher at follow-up at FWE-corrected P<0.05. Results are overlaid on the study-wise QSM template in MNI space, and numbers represent axial slice location in MNI space.

ROI analyses revealed decreasing signed susceptibility over time in Parkinson’s disease in the dentate nucleus (P = 0.032, β = -5.7x10^-5^), red nucleus (P = 0.034, β = -7.4x10^-5^) and insular cortex (P = 0.015, β = -2.2x10^-5^), and increasing signed susceptibility in the medial orbitofrontal cortex (P = 0.045, β = 4.5x10^-5^). However, none of these effects remained significant after correction for multiple comparisons. Of note, we did not find significant signed susceptibility changes over time in the SNpc, SNpr or basal ganglia in patients with Parkinson’s disease. Full ROI results for change in susceptibility over time in Parkinson’s can be seen in Figure 7 and Supplementary Table 3.

**Figure 7.**
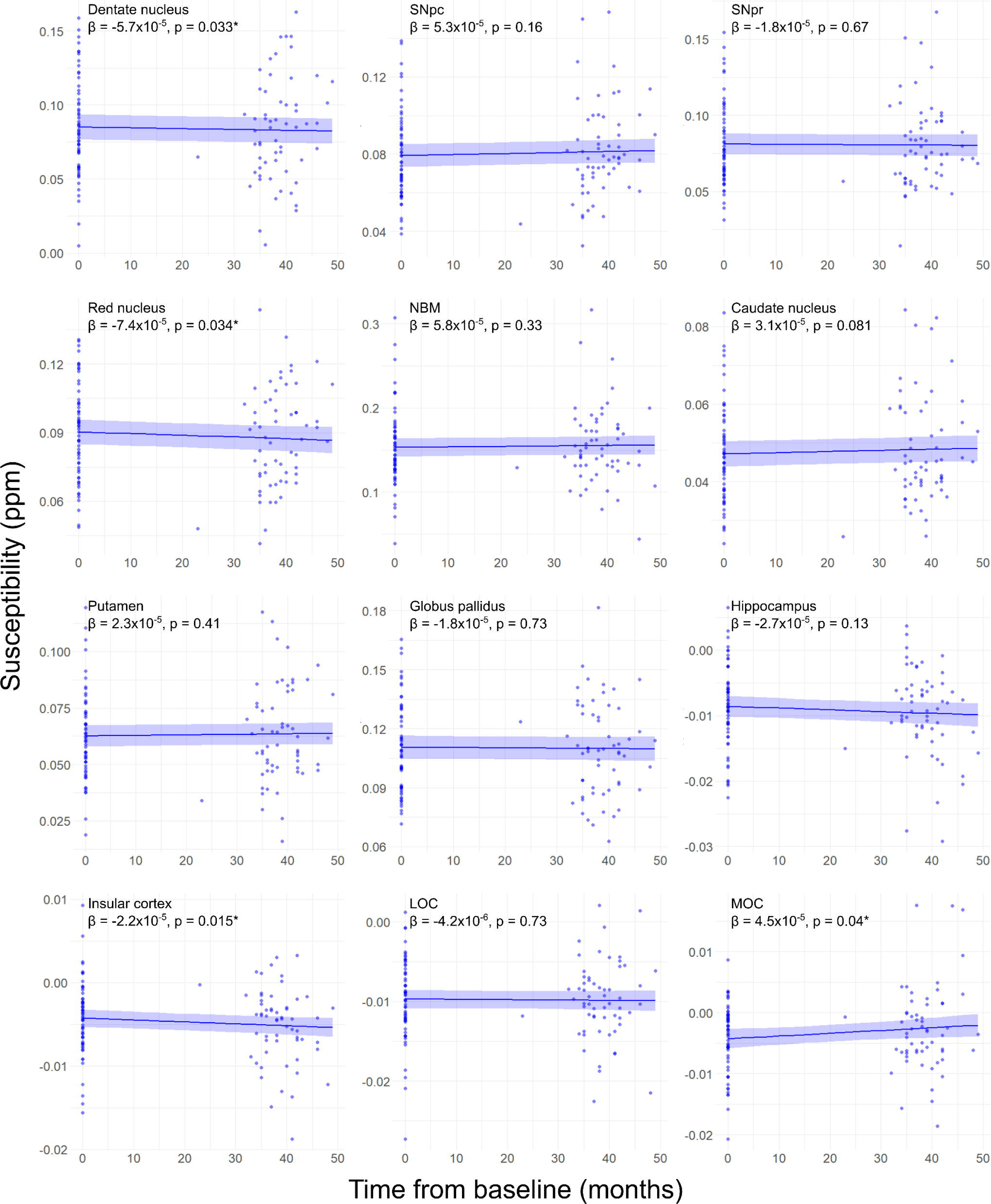
- Results of linear mixed modelling showing regional change in magnetic susceptibility over time in Parkinson’s disease. Change in susceptibility is modelled by fixed effects for time to follow-up (months), age at baseline and sex, and a random intercept effect per subject. β is the coefficient of the fixed effect for time to follow-up on susceptibility. Uncorrected P values indicate the significance of the effect of follow-up time on susceptibility, adjusted for age at baseline, sex and subject. Full ROI statistics can be seen in Supplementary Table 3. SNpc/pr = substantia nigra pars compacta / pars reticulata; NBM = nucleus basalis of Meynert; MOC = medial orbitofrontal cortex; LOC = lateral orbitofrontal cortex.

### Differences in susceptibility between controls and people with Parkinson’s

No significant clusters were found at whole brain level for the comparison of absolute susceptibilities between controls and people with Parkinson’s disease. At uncorrected thresholds (p<0.05), there were increases in absolute susceptibility in the SN and frontal cortices at both baseline and follow-up. At ROI level, in line with previous studies^6, 27, 34–43^, we found that signed magnetic susceptibility was elevated in PD relative to controls at baseline and follow-up in both the SNpc (baseline / follow-up P = 0.025/0.015, T = 2.28/2.48) and SNpr (P = 0.026/0.046 T = 2.27/2.03).

### Voxel based morphometry

We did not find any significant associations between grey or white matter volume at baseline or follow-up and clinical measures at baseline or follow-up in the control or Parkinson’s disease groups at P_FWE_ < 0.05.

## Discussion

In this longitudinal study, we have shown for the first time that QSM shows regional brain susceptibility changes that are predictive of future cognitive and motor outcomes in Parkinson’s disease. Specifically, we showed that baseline increases in magnetic susceptibility values are found in right temporal cortex, right basal forebrain and right putamen, in Parkinson’s patients who showed greater cognitive severity after a mean follow-up of 38 months; and that baseline increases in magnetic susceptibility values bilaterally in the basal ganglia, substantia nigra, red nucleus, insular cortex and dentate nucleus relate to greater motor severity after 3-year follow-up. We further showed that after longitudinal follow-up, relevant brain regions including hippocampus and basal forebrain show higher susceptibilities relating to poorer cognition; and basal ganglia and substantia nigra continue to show increased susceptibilities in relation to greater motor severity, both at whole-brain analysis and in region-of-interest analysis. Whole brain changes relating to clinical measures were not seen when using voxel-based morphometry to measure grey and white matter volume.

We previously demonstrated that susceptibility relates to cognitive and motor severity in Parkinson’s disease^6^. This current study now extends those findings by showing that baseline susceptibility changes can predict later cognitive and motor severity in Parkinson’s disease; and that longitudinally, these associations become stronger, with more widespread and relevant brain regions becoming involved.

Although we found susceptibility increases, reflecting higher levels of brain tissue iron, predicting future cognitive and motor severity, our post-hoc region-of-interest analysis did not show significant susceptibility increases over time within relevant brain regions. There have been very few longitudinal studies using QSM in Parkinson’s disease, with only three recently reported^15–17^ all of which similarly failed to show consistent increases in signed susceptibility values over time. All of these used a region of interest approach, rather than whole brain analysis, and were more focused on motor outcomes. Du et al.^15^ examined 72 Parkinson’s patients and 62 controls at baseline and after 18-month follow-up. Although they showed higher signed susceptibility values in Parkinson’s patients compared with controls, and a relationship between change in signed susceptibility and change in motor UPDRS in the SN pars reticulata, they did not find an increase in signed susceptibility in the SN over time in Parkinson’s patients; and they did not find a relationship between change in signed susceptibility and MoCA scores.

Bergsland et al.^16^ examined signed susceptibility values in the ventral posterior substantia nigra in 18 people with Parkinson’s and 16 controls at baseline and after 3-year follow-up. They found greater susceptibility values in the ventral posterior SN than controls at baseline, with an interaction over time, showing an increase in the Parkinson’s group. Interestingly, although the ventral anterior SN showed higher susceptibility values in Parkinson’s than controls, there was no increase over time. They also did not find an association between baseline SN magnetic susceptibility and either baseline or follow-up motor scores. However, their sample size of only 18 patients most likely lacked power to detect this relationship, and they did not report cognitive severity.

Guan et al.^17^ examined longitudinal regional magnetic susceptibility in 38 Parkinson’s patients with mean follow-up of 16.8 months. Notably, patients were relatively younger than our sample (mean 59.8 years compared to 64.6 years at baseline), although with comparable disease duration (mean 3.9 years compared to 4.1 years at baseline). UPDRS-III scores were also found to decrease over the course of the study (mean 24.2 / 19.2 at baseline / follow-up), in comparison to our sample (mean 22.3 / 28.7 at baseline / follow-up). They found increased signed tissue susceptibility in substantia nigra compared with controls, although susceptibility was lower in the caudate nucleus in Parkinson’s patients than controls. They found no correlations between signed tissue susceptibility and motor scores; and they found decreased signed tissue susceptibility in the substantia nigra, globus pallidus and red nucleus after a mean of 16 months follow-up.

By using a whole brain approach and a more sensitive measure of cognitive severity, we were able to reveal that magnetic susceptibility can predict cognitive as well as motor severity, and that stronger relationships between clinical severity are seen after 3 years of progression. However, consistent with these previous reports, we also did not find robust regional increases in susceptibility over time, suggesting that susceptibility may be more limited in its ability to track (than predict) disease changes over time. The small number of longitudinal studies using QSM in Parkinson’s disease may also reflect this lack of an effect, and a publication bias with lack of publication of these negative findings.

Increases in tissue iron promote production of toxic free reactive oxygen species, that in turn cause damage to DNA^44^; affect mitochondrial function^11^; and can modify proteins though reactive aldehydes^45^, together leading to iron-mediated cell death, or ferroptosis^46^. In addition, excess iron can also promote the aggregation of alpha synuclein fibrils^12^. Higher susceptibility in the basal ganglia and other relevant brain regions in patients with poorer motor severity and predicting poorer motor and cognitive outcomes is supportive of iron having a causative role in progressive neurodegeneration in these brain regions. However, the lack of increase in regional susceptibility values over time, suggests that this relationship is likely to be more complex than simply a gross increase in tissue iron over time.

There are several possible reasons to account for this apparent discrepancy: of a relationship between clinical severity and magnetic susceptibility, but the lack of susceptibility increases (in the same regions) over time. One possible reason is that, although iron stored in ferritin macromolecules is by far the greatest contributor to brain tissue susceptibility^47, 48^, there are other factors that contribute to tissue magnetic susceptibility. For example, other metals such as copper, magnesium or calcium have a diamagnetic effect, meaning they reduce overall magnetic susceptibility (relative to water or soft tissue)^49^. Higher levels of myelin will also act to decrease the magnetic susceptibility of tissue^50^. Previous work has demonstrated increased cortical myelin production by oligodendrocytes in PD motor cortex, potentially due to changes in neuronal excitability^51^, if this holds true for other regions then such effects would be expected to counteract the effect of cortical iron change on susceptibility to some extent. Susceptibility can also be affected by changes in the orientation of tissue microstructure^52^. Pathological proteins such as tau, beta-amyloid or alpha-synuclein may also influence overall tissue magnetic susceptibility. In general, proteins tend to be diamagnetic as they contain large numbers of paired electrons^53^ and this would be expected to lower the overall susceptibility. The diamagnetic nature of tau and amyloid have been confirmed in vitro and in a mouse model of AD^54^. A post-mortem human study in human Alzheimer’s and primary age-related tauopathy also demonstrated correlations between diamagnetic susceptibility and tau and amyloid beta concentrations in hippocampal subfields^55^. Finally, tissue ferritin macromolecules may become redistributed after cell death, drawing away major sources of tissue iron, and in this way, lowering tissue magnetic susceptibility.

Another consideration is that whilst the greatest contribution to tissue magnetic susceptibility is tissue iron (and this is reflected in the relationship between susceptibility values and clinical severity), we found no evidence for extensive changes in tissue iron over time. Instead, other factors, with opposing effects on magnetic susceptibility may come into play. These could include higher levels of diamagnetic proteins, higher relative tissue myelin, and neuronal death, all of which could lower brain tissue susceptibility over time. For this reason, robust changes in magnetic susceptibility are not seen when examined regionally over time, even after 3-year follow-up. Previous work examining brain iron accumulation over the lifetime indicates a slow increase over decades^56–61^, with some suggestion that the steepest increases are seen prior to 40 years of age, well before Parkinson’s disease would be expected to manifest^62^. It is possible that by the time PD clinically presents, much of the disease-related iron accumulation has already occurred. Prodromal changes in proteins responsible for metal homeostasis could explain this, and we recently showed that areas with increased iron in PD have a higher intrinsic expression of genes relating to such processes^14^.

Separately examining absolute and signed susceptibility values goes some way to disambiguating this, as negative values reflect diamagnetic susceptibility sources within tissue. Although we were not able to use this approach in our whole-brain analyses due to the smoothing required, our region-of-interest analyses corroborated the pattern of involvement seen in the whole-brain analysis. In almost all regions, the same relationships were seen for the signed and the absolute susceptibility values. In deep brain nuclei including the substantia nigra, basal ganglia, dentate nucleus, red nucleus, and NBM, ROI analyses using signed versus absolute susceptibilities were near identical for the relevant significant relationships. However, in hippocampal and cortical regions, the direction of the statistical relationships diverged, with increased absolute susceptibility but *decreased* signed susceptibility related to poorer cognitive and motor performance. This suggests that relationships in these regions may include a contribution from decreasing negative (i.e. increasingly diamagnetic) susceptibility.

In future, sequences sensitive to other tissue measures, such as multiparameter maps, with sensitivity to myelin, or alternative imaging with complementary information such as amyloid PET-CT, could be used alongside QSM to provide this additional information. Ultimately, combining in-vivo then ex-vivo MRI with post-mortem histology will enable us to fully disambiguate changes on MRI with tissue composition to provide a more complete picture of neurodegenerative changes in Parkinson’s dementia.

The neuroanatomical locations showing increased magnetic susceptibility with cognitive severity in left precentral gyrus, left middle frontal cortex and right middle temporal gyrus are consistent with previous regional changes found by other groups using different modalities. For example, reductions in grey-matter volume and cortical thickness have been previously reported in the temporal^63–68^, parietal^63, 64, 66, 68, 69^ and frontal^63–67, 70^ cortices in longitudinal studies of Parkinson’s disease.

Notably, in our whole-brain analysis, increased baseline susceptibility in the right NBM was associated with poorer cognitive performance at follow-up; with additional involvement of bilateral hippocampi, temporal cortex, red nucleus and orbitofrontal cortices. The NBM is the major source of cholinergic innervation to the cerebral cortex^71^ and shows post-mortem cell depletion and Lewy-related pathology in PD dementia^72^. Previous neuroimaging has shown decreased NBM volume and increased NBM mean diffusivity in PD with cognitive impairment versus without, and additionally found that this is predictive of developing cognitive impairment in cognitively intact PD^73^. NBM volume is also associated with cortical thickness in PD with mild cognitive impairment^74^, as well as reduced EEG alpha activity in Lewy-body dementia as compared to healthy controls and patients with Alzheimer’s disease^75^. Our finding of increased magnetic susceptibility in this region, relating to cognitive decline after 3-year follow-up, highlights the NBM as a relevant region of early involvement in Parkinson’s dementia.

The hippocampus has also been previously implicated in Parkinson’s with cognitive involvement, for example in a previous meta-analysis implicating functional networks centred on the hippocampus^76^. Decreasing hippocampal volume over time in PD is associated with impaired verbal learning^77^ and may also be predictive of progression to dementia^78^. Hippocampal volume loss or cortical thinning over time is also described in Parkinson’s with mild cognitive impairment^77–81^; and increased hippocampal signed susceptibility is associated with poorer cognitive ability in Parkinson’s disease in previous ROI analyses^7, 8^. Similarly, the orbitofrontal cortex has also been implicated, with longitudinal thinning of the orbitofrontal cortex reported in Parkinson’s disease with mild cognitive impairment^79^, and increased signed susceptibility in the orbitofrontal cortex described in PD with mild cognitive impairment^7^. However, the current analysis indicated a reverse relationship, with poorer cognition associated with decreasing signed susceptibility in these regions. This may be attributable to differences in susceptibility map calculation, or in cohort selection, as our association was found across PD in general, rather than specifically in PD with MCI. Again, future additional work incorporating sequences such as multiparameter maps and amyloid and tau PET could investigate the reasons for these discrepancies.

### Limitations

We did not find significant differences in magnetic susceptibility values between controls and people with Parkinson’s disease in the whole-brain level analysis at either timepoint, after correction for multiple comparisons. On lowering the statistical threshold, increased absolute susceptibility was seen in Parkinson’s patients in the substantia nigra at both timepoints, and we also observed this at ROI level, consistent with many previous studies^33^. Reducing the threshold also indicated increased absolute susceptibility in the frontal cortex at baseline and follow-up, consistent with our previous work and other studies investigating cortical grey matter in PD^6, 27^. The current study was powered to detect susceptibility changes over time in people with Parkinson’s disease, rather than for comparing susceptibility changes between PD and control groups, and our lack of power likely reflects the relatively small control group in this longitudinal study. Our findings so far can only relate to Parkinson’s patients examined as a group. Ultimately, imaging changes providing information on an individual level will be needed to provide prognostic information to individuals in the clinic.

### Conclusions

In summary, we have shown that magnetic susceptibility changes in relevant brain regions predict cognitive and motor severity after 3-year follow-up; with increased susceptibility in the Nucleus Basalis of Meynert related to poorer future cognitive severity; and increased magnetic susceptibility in the basal ganglia related to poorer future motor severity. These findings were seen in both whole-brain and region-of-interest analyses. At follow-up, these relationships between regional magnetic susceptibility and cognitive and motor severity were strengthened, although susceptibility increases were not found within individual brain regions over time. Future work should combine imaging modalities sensitive to myelin, as well as pathological protein accumulation, to shed additional light on the specific tissue changes seen in Parkinson’s dementia and how these relate to measures of clinical severity.

## Data Availability

Code for running group-level analysis will be available upon full publication of the manuscript. Anonymised group-level data may be available upon request.

## Acknowledgements

We are grateful to the participants of this study for volunteering their time and their enthusiastic support for this work.

## Abbreviations

NBM: nucleus basalis of Meynert;
QSM: quantitative susceptibility mapping;
SN: substantia nigra;
SNpc: substantia nigra pars compacta;
SNpr: substantia nigra pars reticulata;
VBM: voxel based morphometry;

