## Supplementary for "Magnetic susceptibility is predictive of future clinical severity in Parkinson’s disease"

**Supplementary tables 1-3**

**Supplementary figures 1-6**

| | Baseline susceptibility vs baseline composite cognitive score | | | Baseline susceptibility vs follow-up composite cognitive score | | | Follow-up susceptibility vs follow-up composite cognitive score | | |
|---|---|---|---|---|---|---|---|---|---|
| | **β** | **P** | **$P_{FDR}$** | **β** | **P** | **$P_{FDR}$** | **β** | **P** | **$P_{FDR}$** |
| *Absolute susceptibility* | | | | | | | | | |
| Dentate nucl. | -9.4e-03 | 0.12 | 0.21 | -9.0e-03 | 0.063 | 0.083 | -9.7e-03 | 0.054 | 0.065 |
| SNpc | -4.7e-03 | 0.27 | 0.30 | -6.3e-03 | 0.056 | 0.083 | ***-9.2e-03*** | ***0.0098*** | ***0.015*** |
| SNpr | -6.0e-03 | 0.21 | 0.29 | ***-9.6e-03*** | ***0.013*** | ***0.032*** | ***-8.3e-03*** | ***0.035*** | ***0.047*** |
| Red nucleus | -9.0e-03 | 0.023 | 0.068 | ***-7.8e-03*** | ***0.0088*** | ***0.026*** | ***-1.0e-02*** | ***0.0013*** | ***0.0079*** |
| NBM | -1.0e-02 | 0.20 | 0.29 | ***-1.8e-02*** | ***0.0039*** | ***0.024*** | ***-1.6e-02*** | ***0.0082*** | ***0.015*** |
| Caudate nucl. | -5.4e-03 | 0.019 | 0.068 | ***-5.3e-03*** | ***0.0038*** | ***0.024*** | ***-5.4e-03*** | ***0.0061*** | ***0.015*** |
| Putamen | -6.5e-03 | 0.055 | 0.13 | ***-7.3e-03*** | ***0.0067*** | ***0.026*** | -5.1e-03 | 0.080 | 0.087 |
| Globus pallidus | -4.3e-03 | 0.30 | 0.30 | -6.0e-03 | 0.077 | 0.084 | -3.4e-03 | 0.32 | 0.32 |
| Hippocampus | -1.4e-03 | 0.074 | 0.15 | -1.3e-03 | 0.036 | 0.072 | ***-1.9e-03*** | ***0.0052*** | ***0.015*** |
| Insular cortex | -1.1e-03 | 0.014 | 0.068 | -6.9e-04 | 0.050 | 0.083 | ***-1.2e-03*** | ***0.0019*** | ***0.0079*** |
| LOC | ***-2.2e-03*** | ***0.0024*** | ***0.029*** | -1.1e-03 | 0.069 | 0.083 | ***-1.7e-03*** | ***0.0056*** | ***0.015*** |
| MOC | -1.0e-03 | 0.28 | 0.30 | -5.4e-05 | 0.94 | 0.94 | ***-2.0e-03*** | ***0.0087*** | ***0.015*** |
| *Signed susceptibility* | | | | | | | | | |
| Dentate nucl. | -9.4e-03 | 0.13 | 0.31 | -9.0e-03 | 0.068 | 0.12 | -9.7e-03 | 0.061 | 0.082 |
| SNpc | -4.6e-03 | 0.29 | 0.44 | -6.2e-03 | 0.065 | 0.12 | ***-9.4e-03*** | ***0.011*** | ***0.030*** |
| SNpr | -6.0e-03 | 0.25 | 0.44 | ***-1.0e-02*** | ***0.013*** | ***0.032*** | *-8.7e-03* | *0.041* | *0.065* |
| Red nucleus | -8.9e-03 | 0.024 | 0.14 | ***-7.8e-03*** | ***0.0092*** | ***0.028*** | ***-1.0e-02*** | ***0.0014*** | ***0.016*** |
| NBM | -9.0e-03 | 0.29 | 0.44 | ***-1.8e-02*** | ***0.0076*** | ***0.028*** | ***-1.6e-02*** | ***0.013*** | ***0.030*** |
| Caudate nucl. | -5.4e-03 | 0.023 | 0.14 | ***-5.4e-03*** | ***0.0044*** | ***0.028*** | ***-5.5e-03*** | ***0.0066*** | ***0.030*** |
| Putamen | -6.4e-03 | 0.062 | 0.19 | ***-7.4e-03*** | ***0.0068*** | ***0.028*** | -5.3e-03 | 0.074 | 0.089 |
| Globus pallidus | -3.0e-03 | 0.50 | 0.55 | -5.4e-03 | 0.134 | 0.20 | -3.1e-03 | 0.39 | 0.39 |
| Hippocampus | -8.1e-04 | 0.50 | 0.55 | 2.5e-04 | 0.80 | 0.80 | 2.0e-03 | 0.043 | 0.065 |
| Insular cortex | -7.5e-05 | 0.93 | 0.93 | 3.2e-04 | 0.61 | 0.66 | ***1.4e-03*** | ***0.023*** | ***0.046*** |
| LOC | 1.7e-03 | 0.059 | 0.19 | 7.9e-04 | 0.30 | 0.39 | ***1.7e-03*** | ***0.012*** | ***0.030*** |
| MOC | 8.2e-04 | 0.46 | 0.55 | 7.2e-04 | 0.40 | 0.49 | 1.0e-03 | 0.34 | 0.36 |

Supplementary Table 1 – Regional associations between signed and absolute susceptibility and cognitive ability assessed using a composite cognitive score. Baseline vs baseline results are adjusted for age at baseline and sex. Baseline vs follow-up results are adjusted for age at baseline, sex and time between scans. Follow-up vs follow-up results are adjusted for age at follow-up and sex. Uncorrected and FDR-corrected p-values are presented, with bold typeface indicating significant associations. β is the linear model coefficient associated with composite cognitive score. SNpc/pr = substantia nigra pars compacta / pars reticulata; NBM = nucleus basalis of Meynert; MOC = medial orbitofrontal cortex; LOC = lateral orbitofrontal cortex.

| | Baseline susceptibility vs baseline UPDRS-III | | | Baseline susceptibility vs follow-up UPDRS-III | | | Follow-up susceptibility vs follow-up UPDRS-III | | |
|---|---|---|---|---|---|---|---|---|---|
| | **β** | **P** | **$P_{FDR}$** | **β** | **P** | **$P_{FDR}$** | **β** | **P** | **$P_{FDR}$** |
| *Absolute susceptibility* | | | | | | | | | |
| Dentate nucl. | 3.0e-04 | 0.41 | 0.61 | ***1.6e-03*** | ***8.8e-4*** | ***0.011*** | ***1.6e-03*** | ***0.0018*** | ***0.013*** |
| SNpc | 3.5e-04 | 0.15 | 0.55 | *6.8e-04* | *0.044* | *0.062* | 6.4e-04 | 0.084 | 0.11 |
| SNpr | 5.0e-04 | 0.074 | 0.45 | ***9.4e-04*** | ***0.018*** | ***0.031*** | ***1.2e-03*** | ***0.0022*** | ***0.013*** |
| Red nucleus | 2.3e-04 | 0.32 | 0.55 | ***9.3e-04*** | ***0.0020*** | ***0.012*** | *6.9e-04* | *0.042* | *0.063* |
| NBM | 8.3e-04 | 0.074 | 0.45 | ***1.9e-03*** | ***0.0029*** | ***0.012*** | ***1.8e-03*** | ***0.0046*** | ***0.019*** |
| Caudate nucl. | 1.5e-04 | 0.25 | 0.55 | ***4.6e-04*** | ***0.015*** | ***0.031*** | ***4.7e-04*** | ***0.020*** | ***0.036*** |
| Putamen | 9.9e-05 | 0.62 | 0.72 | ***7.2e-04*** | ***0.0088*** | ***0.027*** | ***7.5e-04*** | ***0.010*** | ***0.031*** |
| Globus pallidus | 2.8e-04 | 0.25 | 0.55 | ***8.2e-04*** | ***0.016*** | ***0.031*** | ***7.9e-04*** | ***0.021*** | ***0.036*** |
| Hippocampus | 2.0e-05 | 0.66 | 0.72 | -3.1e-05 | 0.62 | 0.62 | 1.2e-04 | 0.11 | 0.13 |
| Insular cortex | -1.5e-05 | 0.57 | 0.72 | *7.2e-05* | *0.047* | *0.062* | ***9.2e-05*** | ***0.019*** | ***0.036*** |
| LOC | -9.8e-06 | 0.82 | 0.82 | 9.7e-05 | 0.10 | 0.12 | 4.3e-05 | 0.50 | 0.54 |
| MOC | -5.7e-05 | 0.30 | 0.55 | 4.1e-05 | 0.60 | 0.62 | 3.6e-05 | 0.65 | 0.65 |
| *Signed susceptibility* | | | | | | | | | |
| Dentate nucl. | 3.2e-04 | 0.38 | 0.51 | ***1.6e-03*** | ***8.2e-4*** | ***0.010*** | ***1.6e-03*** | ***0.0017*** | ***0.014*** |
| SNpc | 3.6e-04 | 0.15 | 0.51 | 6.8e-04 | 0.050 | 0.067 | 6.4e-04 | 0.094 | 0.13 |
| SNpr | 5.4e-04 | 0.073 | 0.44 | ***9.8e-04*** | ***0.021*** | ***0.034*** | ***1.3e-03*** | ***0.0023*** | ***0.014*** |
| Red nucleus | 2.4e-04 | 0.31 | 0.51 | ***9.4e-04*** | ***0.0018*** | ***0.011*** | *7.0e-04* | *0.042* | *0.063* |
| NBM | 9.0e-04 | 0.065 | 0.44 | ***2.0e-03*** | ***0.0034*** | ***0.014*** | ***1.8e-03*** | ***0.0055*** | ***0.022*** |
| Caudate nucl. | 1.5e-04 | 0.27 | 0.51 | ***4.7e-04*** | ***0.017*** | ***0.033*** | ***4.8e-04*** | ***0.020*** | ***0.041*** |
| Putamen | 1.1e-04 | 0.60 | 0.66 | ***7.2e-04*** | ***0.0096*** | ***0.027*** | ***7.4e-04*** | ***0.013*** | ***0.040*** |
| Globus pallidus | 2.4e-04 | 0.35 | 0.51 | ***7.9e-04*** | ***0.031*** | ***0.046*** | *7.7e-04* | *0.036* | *0.061* |
| Hippocampus | -8.1e-05 | 0.25 | 0.51 | 7.1e-05 | 0.48 | 0.52 | -1.4e-04 | 0.17 | 0.21 |
| Insular cortex | -4.6e-05 | 0.33 | 0.51 | ***-1.6e-04*** | ***0.011*** | ***0.027*** | ***-1.5e-04*** | ***0.021*** | ***0.041*** |
| LOC | 6.2e-07 | 0.99 | 0.99 | -1.9e-05 | 0.80 | 0.80 | -1.3e-05 | 0.86 | 0.86 |
| MOC | 3.4e-05 | 0.58 | 0.66 | -1.6e-04 | 0.071 | 0.085 | -3.4e-05 | 0.76 | 0.83 |

Supplementary Table 2 – Regional associations between signed and absolute susceptibility and motor ability assessed with the UPDRS-III score. Baseline vs baseline results are adjusted for age at baseline and sex. Baseline vs follow-up results are adjusted for age at baseline, sex and time between scans. Follow-up vs follow-up results are adjusted for age at follow-up and sex. Uncorrected and FDR-corrected p-values are presented, with bold typeface indicating significant associations. β is the linear model coefficient associated with UPDRS-III score. SNpc/pr = substantia nigra pars compacta / pars reticulata; NBM = nucleus basalis of Meynert; MOC = medial orbitofrontal cortex; LOC = lateral orbitofrontal cortex.

| | Marginal $R^2$ | Conditional $R^2$ | β | P | $P_{FDR}$ |
|---|---|---|---|---|---|
| Dentate nucleus | 0.064 | 0.97 | $-5.7x10^{-5}$ | 0.033 | 0.13 |
| SNpc | 0.017 | 0.89 | $5.3\ x10^{-5}$ | 0.16 | 0.27 |
| SNpr | 0.036 | 0.90 | $-1.8x10^{-5}$ | 0.68 | 0.73 |
| Red nucleus | 0.022 | 0.89 | $-7.4x10^{-5}$ | 0.034 | 0.13 |
| NBM | 0.16 | 0.93 | $5.8x10^{-5}$ | 0.33 | 0.50 |
| Caudate nucleus | 0.084 | 0.93 | $3.0x10^{-5}$ | 0.081 | 0.20 |
| Putamen | 0.18 | 0.92 | $2.3x10^{-5}$ | 0.41 | 0.55 |
| Globus pallidus | 0.027 | 0.87 | $-1.8x10^{-5}$ | 0.66 | 0.72 |
| Hippocampus | 0.036 | 0.68 | $-2.7x10^{-5}$ | 0.13 | 0.25 |
| Insula | 0.16 | 0.82 | $-2.2x10^{-5}$ | 0.015 | 0.13 |
| LOC | 0.15 | 0.75 | $-4.2x10^{-6}$ | 0.73 | 0.73 |
| MOC | 0.044 | 0.47 | $4.5x10^{-5}$ | 0.045 | 0.13 |

Supplementary Table 3 – Results of linear mixed modelling showing regional change in signed magnetic susceptibility over time in Parkinson's disease. Change in susceptibility is modelled by fixed effects for time to follow-up (months), age at baseline and sex, and a random intercept effect per subject. The marginal $R^2$ indicates the variance explained by fixed effects only, while the conditional $R^2$ indicates variance explained by both fixed and random effects. Β is the coefficient of the fixed effect for time to follow-up on susceptibility, positive values indicate increasing susceptibility over time and negative values indicate decreasing susceptibility over time. Uncorrected P values indicate the significance of the effect of follow-up time on susceptibility, adjusted for age at baseline, sex and subject. $P_{FDR}$ values are corrected across multiple comparisons. SNpc/pr = substantia nigra pars compacta / pars reticulata; NBM = nucleus basalis of Meynert; MOC = medial orbitofrontal cortex; LOC = lateral orbitofrontal cortex.

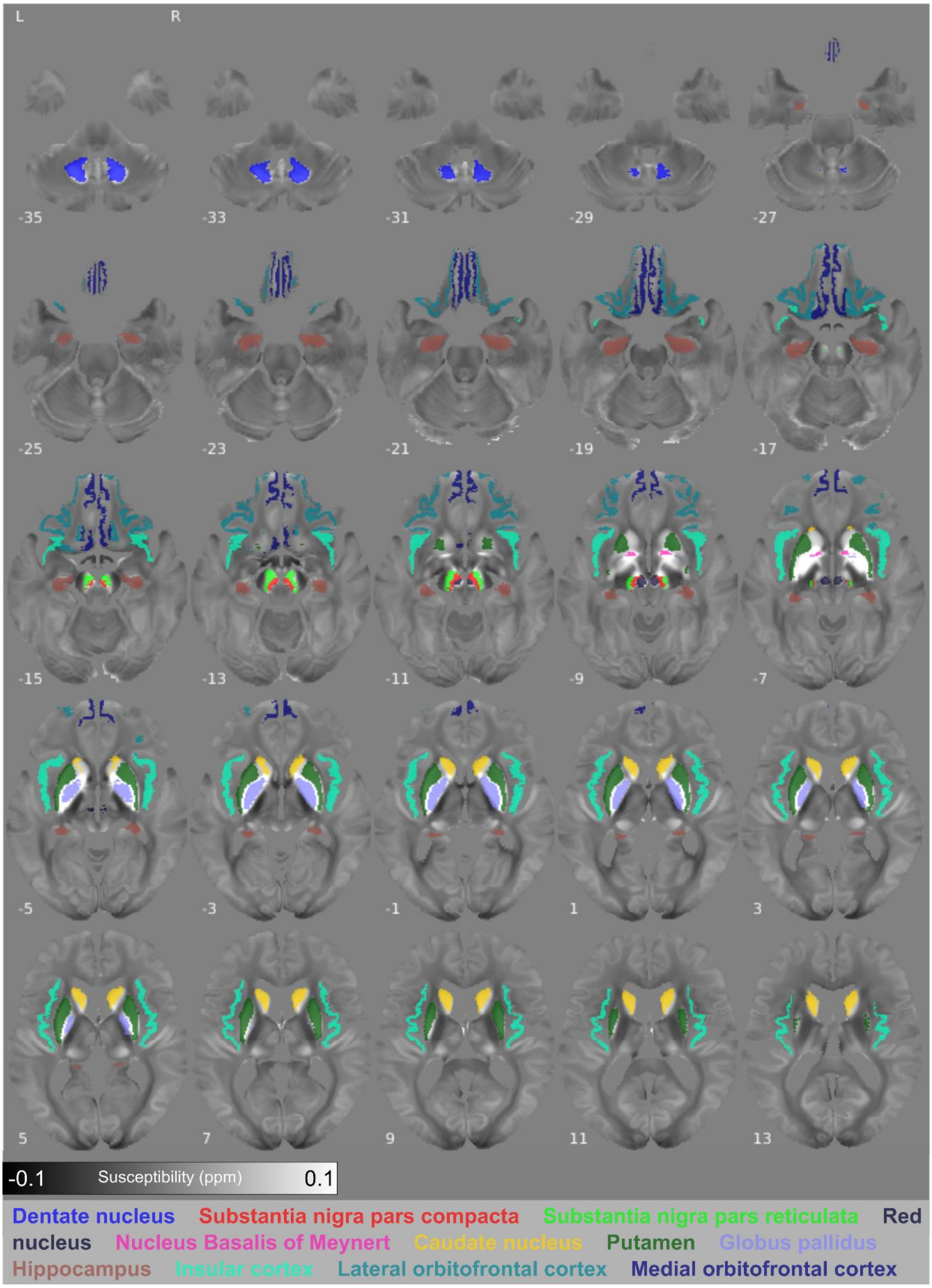


**Supplementary Figure 1 – Regions of interest visualised on the study-wise QSM template in MNI space.** Regions are indicated by the colour coded text in the bottom panel. Numbers indicate axial slice position in MNI space.

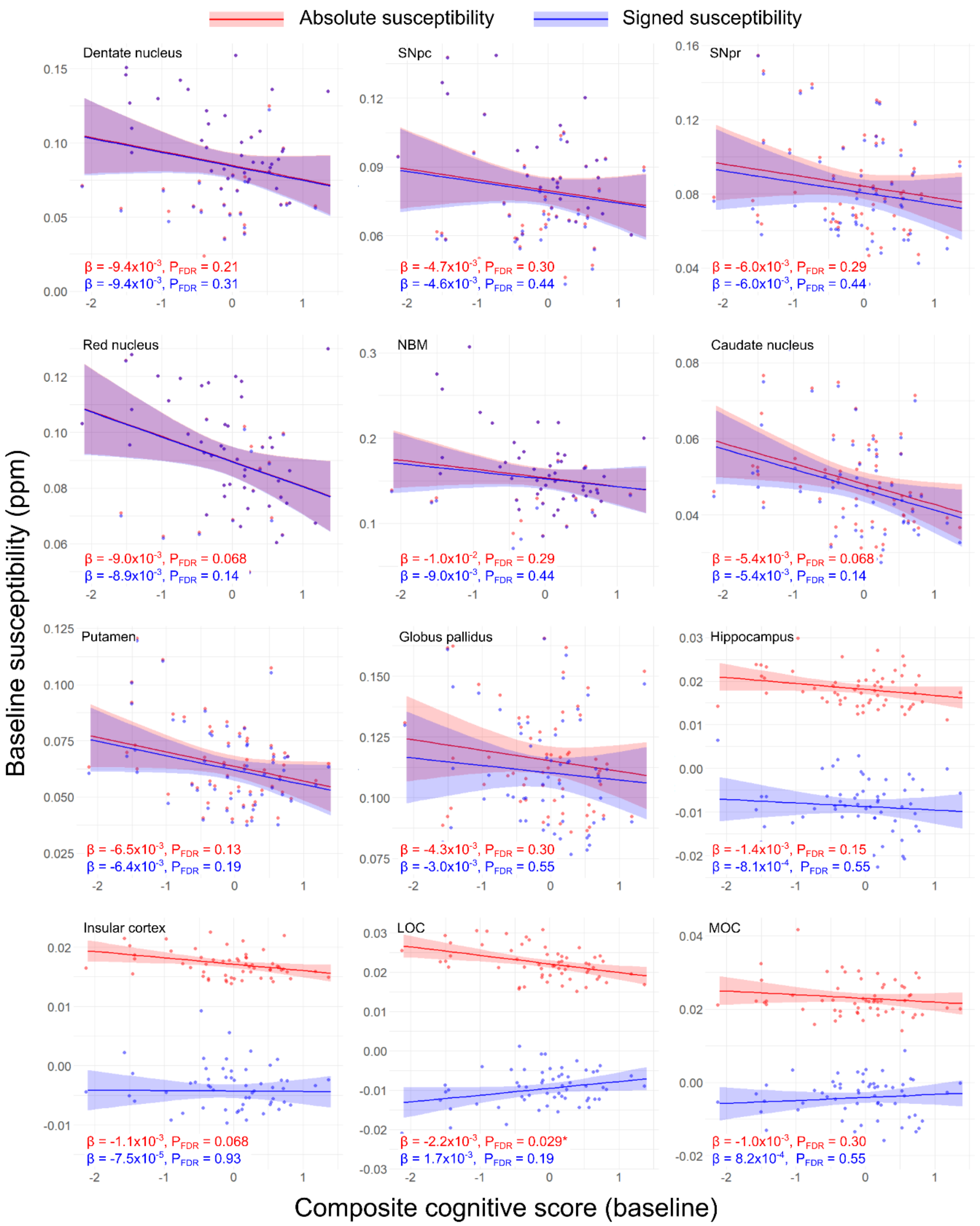


**Supplementary Figure 2 – Regional relationships between baseline magnetic susceptibility and baseline cognitive score in Parkinson's disease.** Data and statistics relating to ROI mean absolute susceptibility are shown in red, and those relating to ROI mean signed susceptibility are shown in blue. Results are adjusted for age at baseline and sex. FDR-corrected p-values ($P_{FDR}$) are presented, with asterisks indicating significant interactions at $P_{FDR} < 0.05$. β is the linear model coefficient associated with composite cognitive score. SNpc/pr = substantia nigra pars compacta / pars reticulata; NBM = nucleus basalis of Meynert; MOC = medial orbitofrontal cortex; LOC = lateral orbitofrontal cortex.

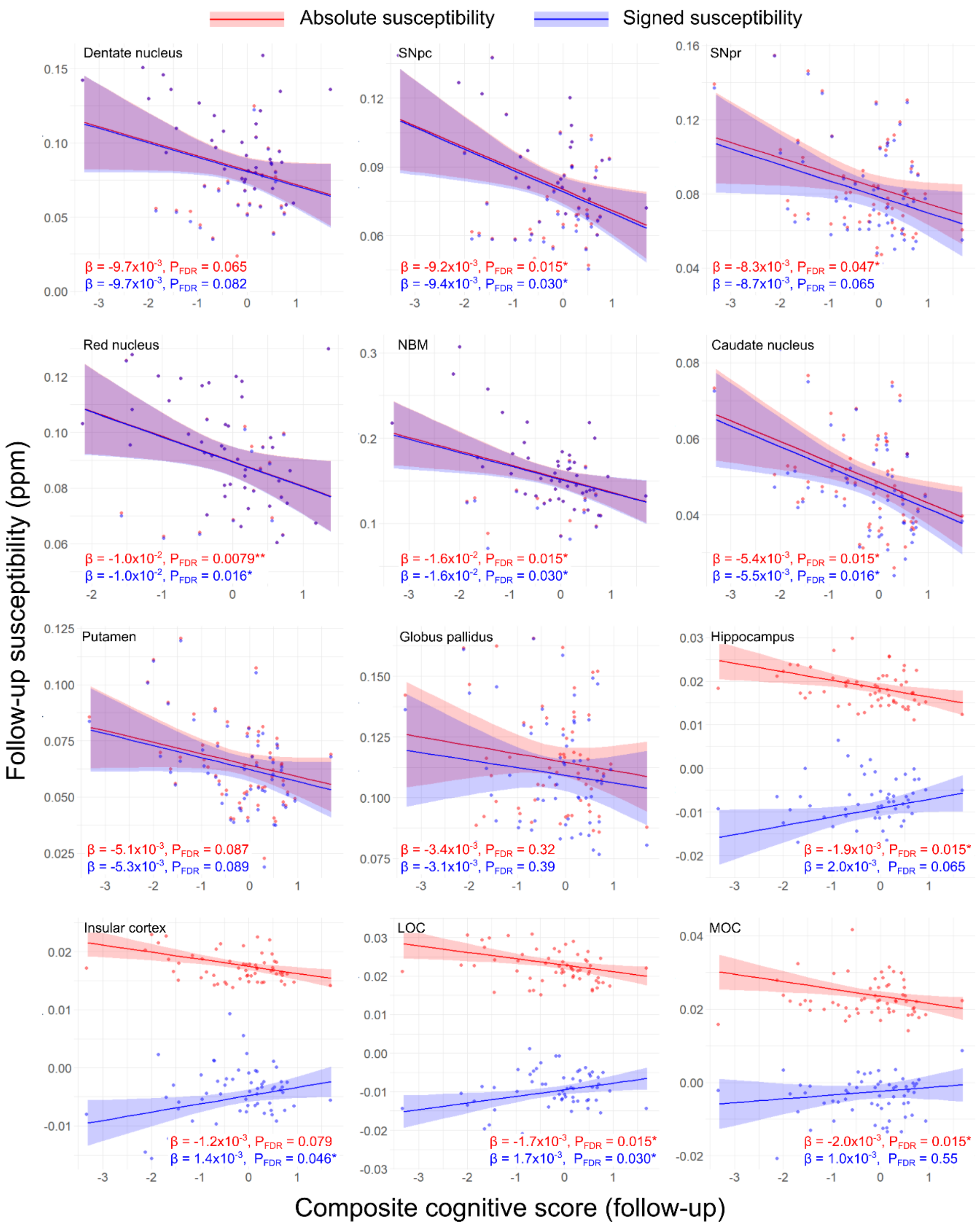


**Supplementary Figure 3 – Regional relationships between follow-up magnetic susceptibility and follow-up cognitive score in Parkinson's disease.** Data and statistics relating to ROI mean absolute susceptibility are shown in red, and those relating to ROI mean signed susceptibility are shown in blue. Results are adjusted for age at follow-up and sex. FDR-corrected p-values ($P_{FDR}$) are presented, with single asterisks indicating significant interactions at $P_{FDR} < 0.05$, and double asterisks indicating $P_{FDR} < 0.01$. β is the linear model coefficient associated with composite cognitive score. SNpc/pr = substantia nigra pars compacta / pars reticulata; NBM = nucleus basalis of Meynert; MOC = medial orbitofrontal cortex; LOC = lateral orbitofrontal cortex.

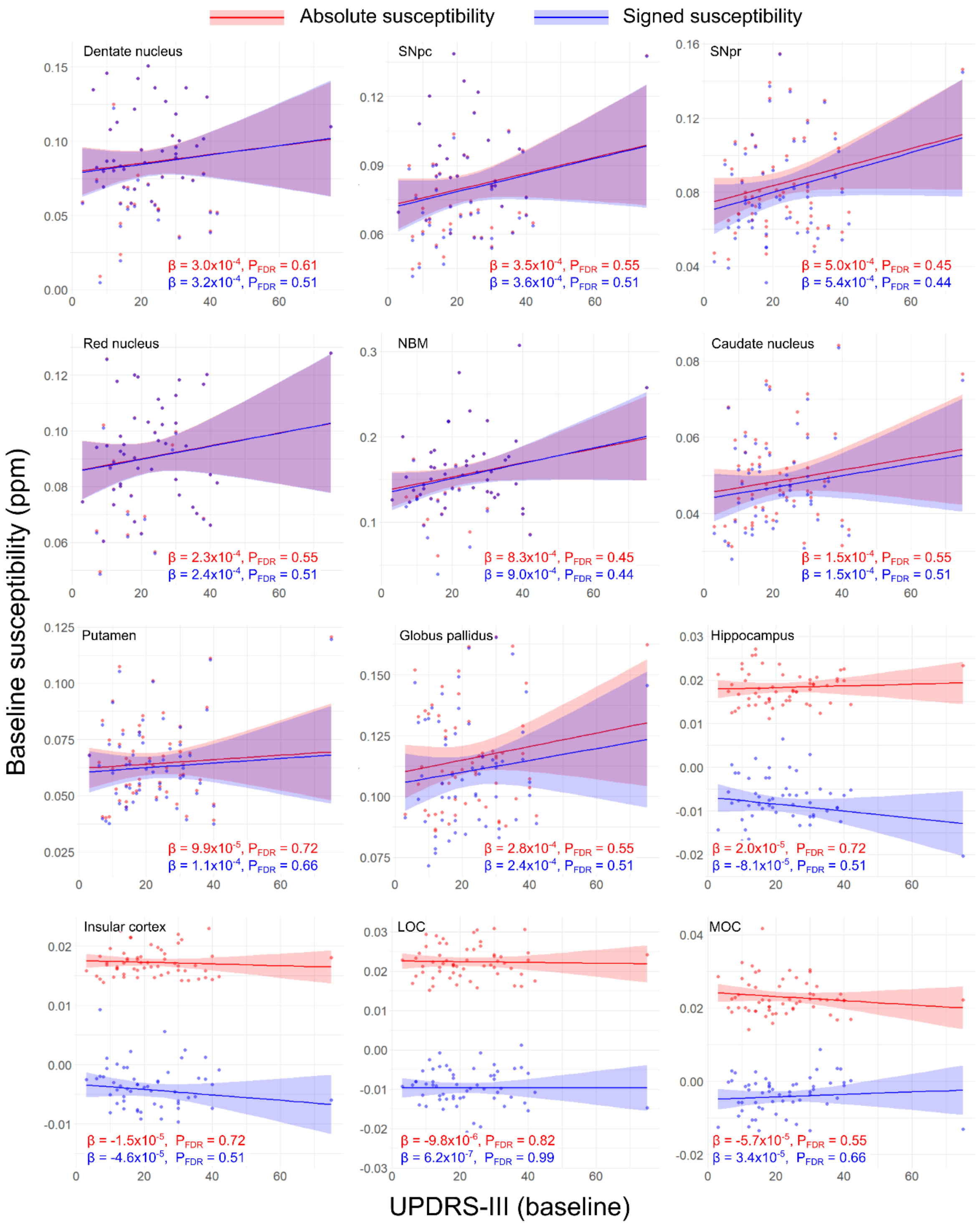


**Supplementary Figure 4 – Regional relationships between baseline magnetic susceptibility and baseline motor severity score in Parkinson's disease.** Data and statistics relating to ROI mean absolute susceptibility are shown in red, and those relating to ROI mean signed susceptibility are shown in blue. Results are adjusted for age at baseline and sex. FDR-corrected p-values ($P_{FDR}$) are presented. β is the linear model coefficient associated with MDS-UPDRS-III score. SNpc/pr = substantia nigra pars compacta / pars reticulata; NBM = nucleus basalis of Meynert; MOC = medial orbitofrontal cortex; LOC = lateral orbitofrontal cortex. MDS-UPDRS-III = Movement Disorders Society Unified Parkinson's Disease Rating Scale part III.

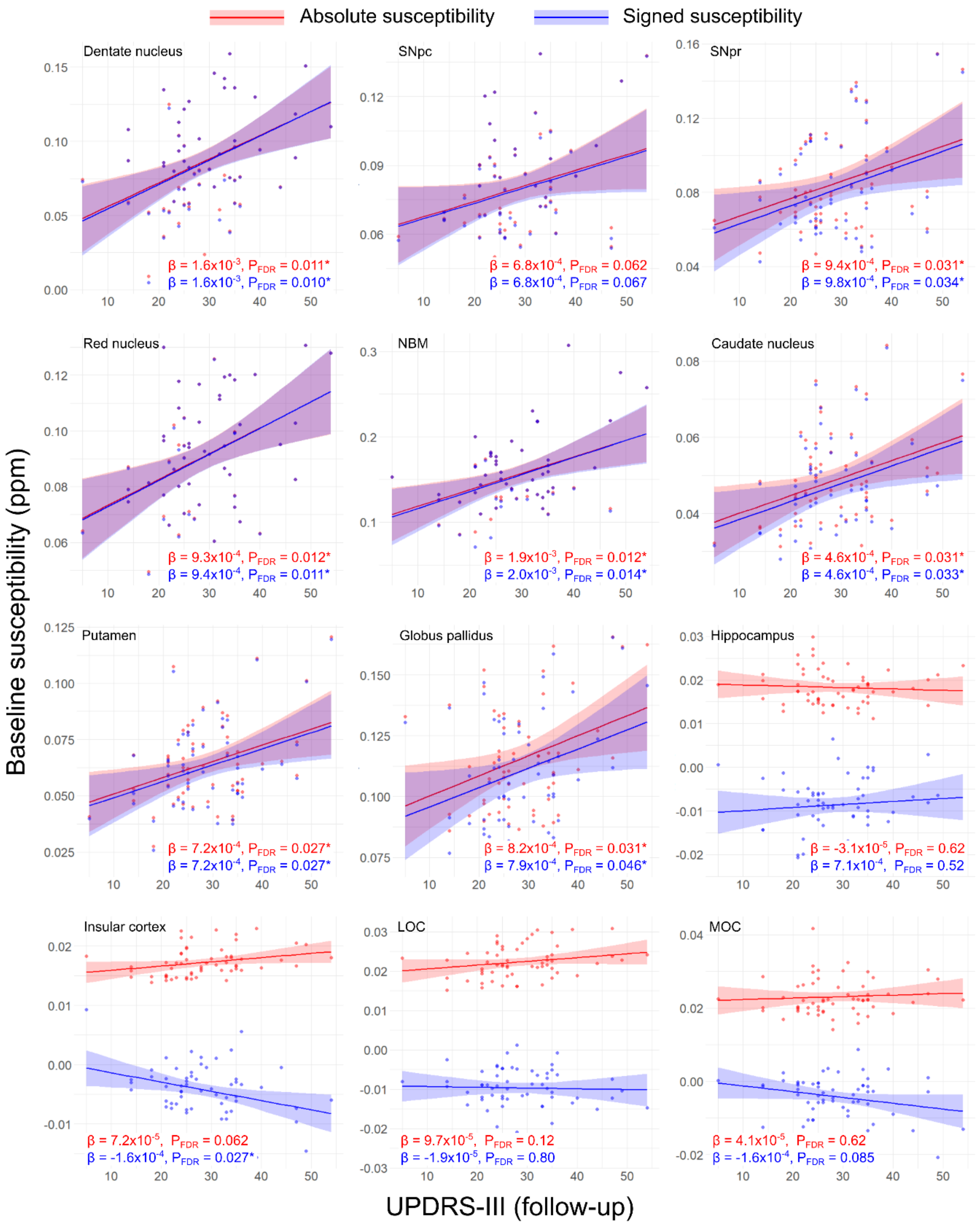


**Supplementary Figure 5 – Regional relationships between follow-up magnetic susceptibility and follow-up motor severity score in Parkinson's disease.** Data and statistics relating to ROI mean absolute susceptibility are shown in red, and those relating to ROI mean signed susceptibility are shown in blue. Results are adjusted for age at follow-up and sex. FDR-corrected p-values ($P_{FDR}$) are presented, with asterisks indicating significant interactions at $P_{FDR} < 0.05$. β is the linear model coefficient associated with MDS-UPDRS-III score. SNpc/pr = substantia nigra pars compacta / pars reticulata; NBM = nucleus basalis of Meynert; MOC = medial orbitofrontal cortex; LOC = lateral orbitofrontal cortex. MDS-UPDRS-III = Movement Disorders Society Unified Parkinson's Disease Rating Scale part III.

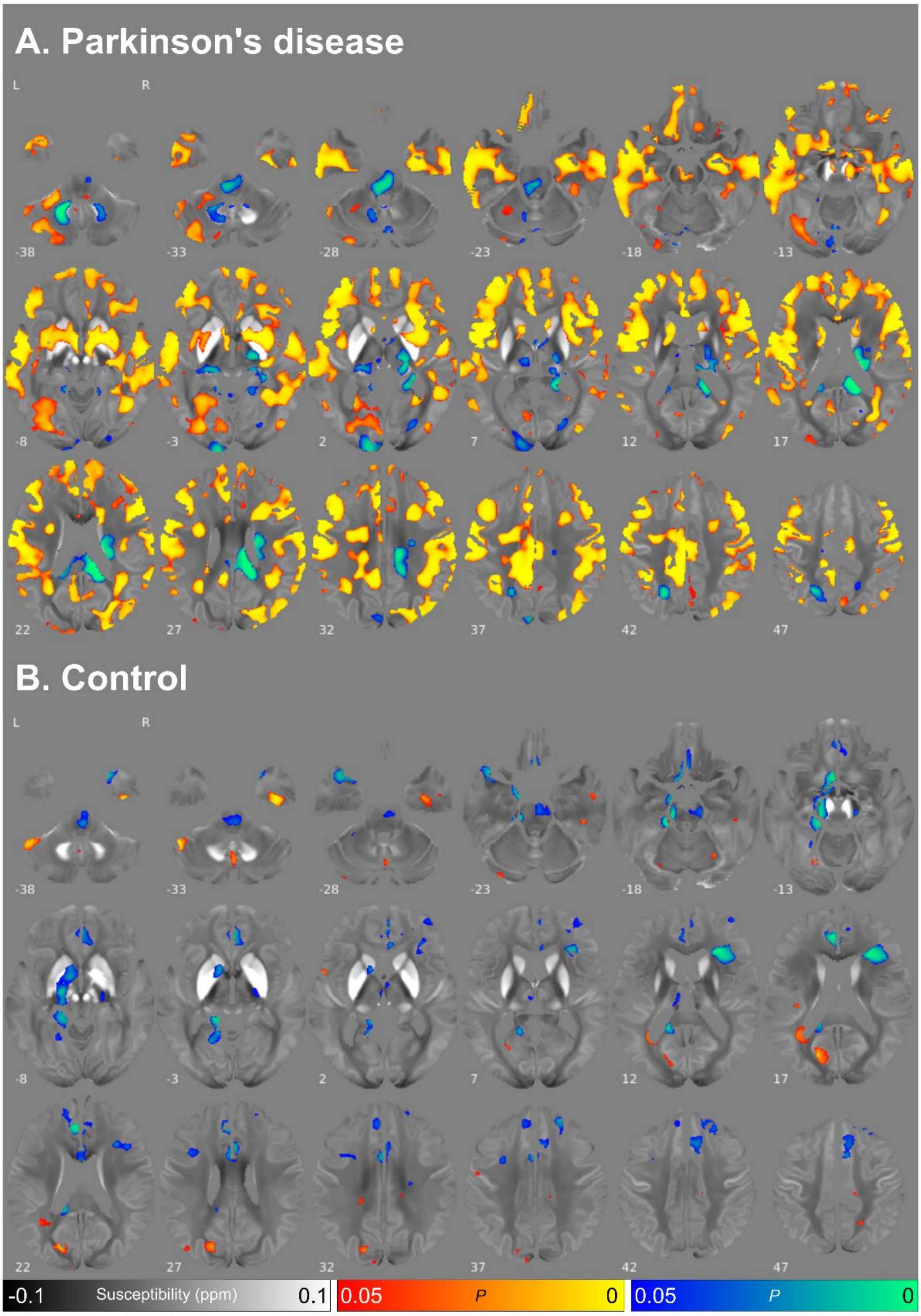


**Supplementary Figure 6 – Changes in absolute magnetic susceptibility over time at whole brain, uncorrected for multiple comparisons.** Results are displayed for **(A)** Parkinson's disease and **(B)** healthy control participants. Analyses are adjusted for age at baseline, sex, and time between scans. Red/yellow clusters represent voxels where absolute QSM was higher at follow-up at uncorrected P<0.05, blue / green clusters represent voxels where absolute QSM was lower at follow-up at uncorrected P<0.05. Results are overlaid on the study-wise QSM template in MNI space, and numbers represent axial slice location in MNI space. Analyses were performed for exploratory purposes.
